# *Ex Vivo* Hypothermic Perfusion Enables 48-Hour Heart Preservation and Bench-Top Functional Recovery via Normothermic Reperfusion in a Porcine Model

**DOI:** 10.1101/2025.09.03.25335053

**Authors:** Chiara Camillo, Morgan K. Moroi, Yaagnik Kosuri, Anthony Campbell, Arianna Adamo, Krushang Patel, Cary Karcher, Sebastian J. Bauer, Diana Albino, Emre Bektik, Ting Peng, Liming Pei, Christine Chan, Kenmond Fung, Miroslav Sekulic, Renu Nandakumar, Erfan Faridmoayer, Claire Kho, Bianca Bernante, Alexander Romanov, Melissa Tamimi, Juan B Grau, Koji Takeda, Giovanni Ferrari

## Abstract

**Background:** *Ex vivo* oxygenated perfusion systems is a promising approach to extend cardiac allograft preservation beyond the typical 4–6h limit allowed by static-cold-storage (SCS). Hypothermic oxygenated perfusion (HOPE) has been proven to safely preserve donor hearts, yet its underlying molecular mechanisms have not been extensively evaluated.

**Objectives:** The aim of the study is to characterize cardiomyocyte viability, transcriptomic and metabolomic responses, and functional recovery of porcine hearts preserved with HOPE for up to 48h, including evaluating their ability to regain sinus rhythm following bench-top normothermic reperfusion (NMP).

**Methods:** Seventeen Yorkshire pigs underwent donor cardiectomy. In the first arm, ten hearts were preserved for up to 48h using either SCS (n=5) or HOPE (n=5). Endomyocardial biopsies were collected at 0, 12, 24, and 48h for histology, RNA sequencing, flow cytometry, and metabolomics. In the second arm, six HOPE-preserved hearts (3h, 24h, 48h) and one SCS-preserved heart (24h) underwent 2h NMP to simulate transplantation and assess reanimation.

**Results:** HOPE preserved cardiomyocyte viability and structural integrity for 48h, in contrast to SCS in both arms of the study. RNA sequencing and untargeted metabolomics revealed conserved energy-substrate profiles in HOPE and progressive ischemic metabolite accumulation in SCS. All HOPE hearts regained stable sinus rhythm.

**Conclusions:** HOPE enables 48h *ex vivo* heart preservation while maintaining cardiomyocyte integrity, normal gross and microscopic architecture, and rapid functional recovery on bench-top reperfusion in a preclinical model. These findings establish a foundation for redefining clinical preservation times and widening geographic donor access.

**GRAPHICAL ABSTRACT:** 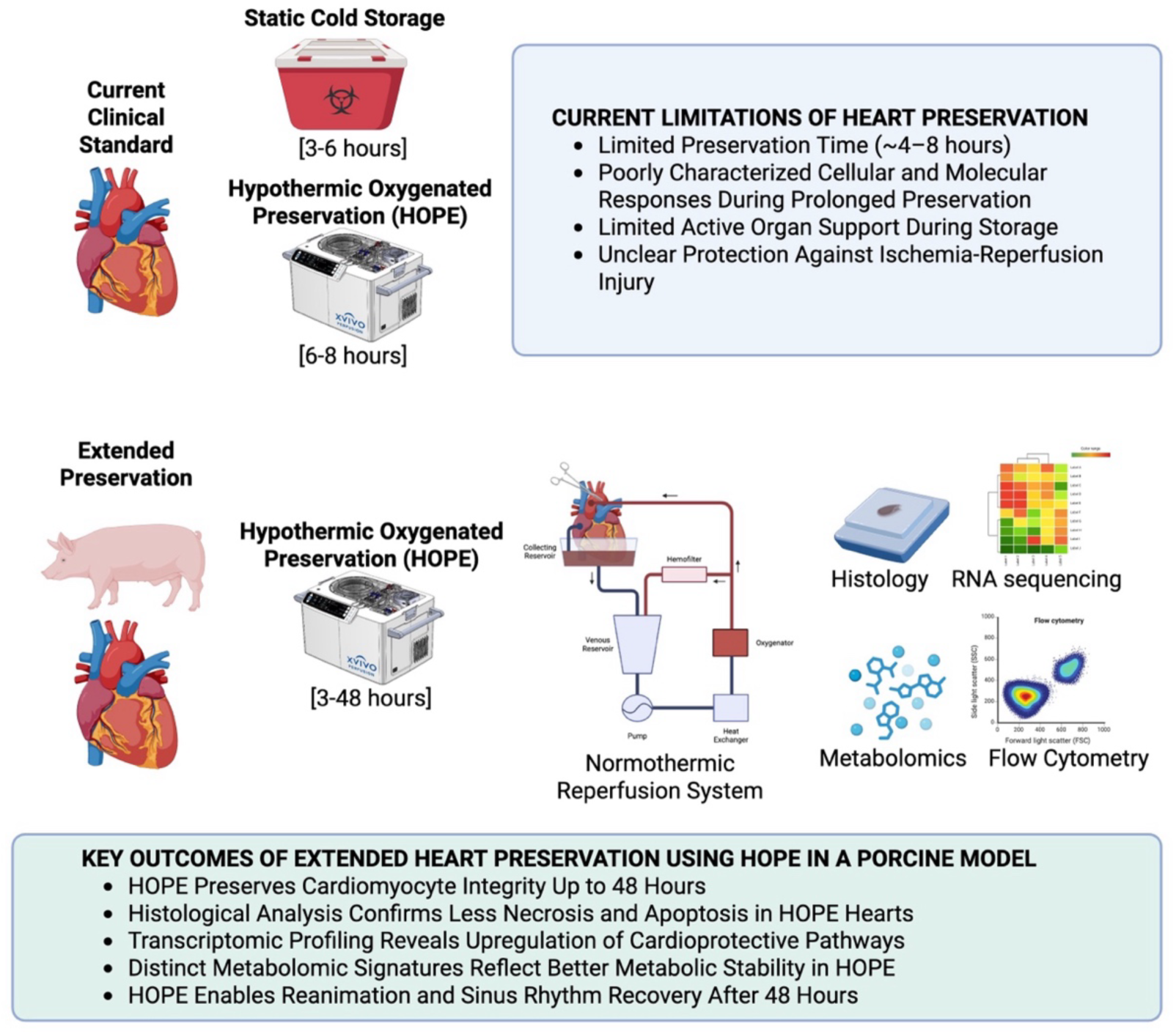

## Introduction

Heart transplantation (HTx) remains the gold standard for the treatment of end-stage heart failure. Despite an increase in the total number of annual heart transplants performed worldwide, with more than 5,000 recorded in 2023 alone, there remains a shortage of donor organs, as nearly 50,000 patients enter the waitlist each year^1^. The ongoing shortage of donor hearts has prompted efforts to expand the donor pool, including the utilization of donation after circulatory death (DCD), expansion of donor criteria, and improvements in organ preservation to allow for procurement from further geographic distances.

For HTx, the standard practice for allograft preservation is static cold storage (SCS), where the graft is maintained in cold saline or cardioplegic solution. Hypothermia is targeted at 0-8°C in order to reduce metabolic activity to approximately 10–12% of the baseline normothermic conditions. Under SCS conditions, ischemic times of more than 4-6 hours have been associated with a higher risk of early graft failure, impaired graft function, and post-transplant mortality^2^. Unfortunately, this short time window serves as a direct geographic barrier, limiting the number of organs available within the donor pool^2,3^.

The recent introduction of *ex vivo* perfusion devices has demonstrated the potential for safe and prolonged storage of cardiac allografts^4,5^. Here, we focus on the use of hypothermic oxygenated perfusion (HOPE) through the XVIVO system, which perfuses the vented heart antegrade using an oxygenated solution via an aortic root cannula. The safety of extended preservation with HOPE for heart transplant has been reported by Steen et al. in 2016^6^. Subsequent HOPE animal studies suggest safe extension of preservation, with See Hoe et al. reporting no cardiac impairment after 8 hours^7^. The first-in-human study using the XVIVO device had a median preservation time of 3 hours and 43 minutes^5^. This was followed by a nonrandomized, single-arm trial that further revealed safe preservation times of up to 8 hours and 47 minutes^8^.

Brouckaert et al. published a randomized, controlled, multicenter clinical trial in 2023^8^. With a median preservation time of 240 min., the Brouckaert et al. trial displayed non-inferiority when comparing XVIVO preservation to SCS regarding the primary endpoint of time-to-first cardiac-related death, acute cellular rejection, primary graft dysfunction, or graft failure. Additionally, case reports showed the feasibility of XVIVO preservation times >12h^9^ and effective utilization of XVIVO heart storage in DCD scenario^10^.

Despite these advances, the maximum duration for safe use of this device remains undefined. Moreover, the cellular and molecular mechanisms enabling prolonged HOPE or other *ex vivo* preservation methods are not fully understood. Finally, the functional performance of hearts after extended HOPE preservation has yet to be thoroughly characterized. This study addresses these gaps by comprehensively evaluating the molecular, cellular, and functional outcomes of porcine hearts preserved with HOPE for up to 48 hours, 6 to 8 times the current standard clinical practice. Using a bench-top normothermic reperfusion system, we also assess the ability of these hearts to regain sinus rhythm, simulating transplant reanimation and establishing an experimental setting for future full retransplantation studies.

## Materials and Methods

### Animal characteristics

Female American Yorkshire pigs (n=17, 45-50 kg, 4-6 months old) were used in both groups, SCS and HOPE. This study was approved by the Columbia University Institutional Animal Care and Use Committee (IACUC) under protocol number AC-AABX9650. The study was performed under the National Institutes of Health (NIH) Guide for the Care and Use of Laboratory Animals.

### Donor Surgical Procedure

On the day of surgery, the swine donor underwent anesthesia induction with intramuscular Telazol (6 mg/kg), propofol (1 mg/kg), and buprenorphine (0.01-0.03 mg/kg). Animals were orally intubated and mechanically ventilated. Anesthesia and analgesia were maintained with inhaled isoflurane and/or continuous infusions of midazolam (0.2 mg/kg/hr) and propofol (3-5 mg/kg/hr). A midline sternotomy was performed with a bone saw, and the heart and great vessels were exposed. Intravenous heparin (300 IU/kg) was given for systemic anticoagulation. The ascending aorta was cannulated with an antegrade cardioplegia cannula (Surge Cardiovascular, Grand Rapids, MI). An aortic cross-clamp was applied, and 2L of cardioplegia at 4°C was delivered. As per our clinical protocols, SCS hearts received 2L of histidine-tryptophan-ketoglutarate (HTK) cardioplegic solution, and HOPE hearts received 2L of XVIVO Heart Solution. Sterile slush was applied to the heart. Following adequate cardiac arrest, a standard cardiectomy was performed. A baseline weight of the heart was acquired, and the heart was prepared for preservation using either SCS or HOPE.

### Preservation Methods

Hearts selected for SCS were stored in 2L of cardioplegic solution at 4°C for up to 48 hours. Hearts selected for HOPE were preserved using the XVIVO Heart Assist Transport (XVIVO Group, Gothenburg, Sweden), which provides oxygenated perfusion using carbogen gas (95% O2, 5% CO2) at 8°C. The heart preservation device was primed with XVIVO cardioplegic solution supplemented with 350ml packed red blood cells and XVIVO heart solution supplement. Preparation of the donor heart for HOPE consisted of aortic cannulation and placement of a silastic tube across the mitral valve to vent the left ventricle. The heart was connected to the device, the device was de-aired, and perfusion was carried out at an aortic root pressure of 20 mmHg for up to 48 hours. Heart weight was recorded again at the end of SCS or XVIVO preservation.

### Bench-Top Normothermic Machine Perfusion

Following standard cardiectomy, six hearts underwent HOPE preservation for 3hr (n=2), 24hrs (n=2), and 48hrs (n=2), followed by a 2-hour period of reperfusion using a normothermic machine perfusion (NMP) circuit, with the intention of simulating reperfusion. One heart underwent SCS preservation for 24hrs followed by reperfusion with NMP. The NMP circuit consisted of a venous reservoir with Capiox FX05 oxygenator (Terumo Cardiovascular, Ann Arbor, MI), 3T Heater-Cooler system (LivaNova, London, UK), and Quantum 4-in. roller pump (Spectrum Medical, Gloucester, UK), and the circuit was primed with 500cc of whole blood from the donor pig. To load the heart onto the circuit, the ascending aorta was cannulated (8Fr Bio-Medicus NextGen Pediatric Arterial Cannula, Medtronic, Minneapolis, MN), and a silastic tube was maintained across the mitral valve to vent the left ventricle. The atria were widely opened, and venous return from the heart was allowed to passively drain into a collecting reservoir. The circuit was slowly initiated, the aorta de-aired, and an aortic cross-clamp was applied to begin 2 hours of reperfusion.

### Contractility Quantification

Stabilized perfusion videos of porcine hearts were analyzed using a custom Python pipeline to quantify myocardial motion. Videos were converted to greyscale, and the frame-to-frame displacements in the horizontal and vertical directions were estimated using the Farnebäck dense optical flow algorithm in OpenCV. Motion magnitude was averaged within a user-defined rectangular region of interest (ROI) and converted from pixels per frame to millimeters per second using the frame rate metadata and a spatial calibration factor derived from a reference marker in the field of view. The velocity signal was smoothed through a Savitzky-Golay filter and local minima, corresponding to end-diastolic phases, were used to ensure average velocity magnitudes were obtained only for full cardiac cycles. If no valid cycles were found, the entire recording was treated as a single cycle and analyzed in full. A detailed protocol is provided in the Supplemental Material and Methods.

### Sampling and Tissue Collection

Throughout the preservation period, 8-mm punch endomyocardial biopsies were obtained from the right ventricular (RV) septum and left ventricular (LV) free wall at 0, 12, 24, and 48 hours for both HOPE-preserved hearts and SCS hearts. Perfusate (10mL) was extracted from the device in K2 EDTA blood collection tubes. Perfusate was centrifuged at 1500 rpm for 15 min at 4°C. The plasma was collected, aliquoted, and stored at -80°C for further analysis. Each tissue was stored concurrently with different preservation methods, such as fixation in formalin, embedded in OCT, and flash frozen in liquid nitrogen for further assay purposes.

### Histological Analysis

Tissue was obtained from the RV septum and LV free wall using an 8-mm biopsy punch at designated time points. Specimens were fixed in 10% formalin for 48 hours, transferred to ethanol 70% solution, and embedded in paraffin. 5µm-thick sections were cut via microtome and mounted on high-adherence glass microscope slides. Histologic sections of tissue were stained with hematoxylin and eosin (H&E) as well as Masson’s trichrome and visualized on a Leica 10X microscope. Standard TUNEL staining was also performed to assess apoptotic cells.

### Flow Cytometry

Fresh porcine cardiac tissue (0.2–0.3 g) was collected from the right ventricular septum using an 8 mm biopsy punch at designated time points. Tissue was enzymatically digested in a collagenase/DNase/hyaluronidase solution and dissociated into single-cell suspensions. Following red blood cell lysis and fixation, cells were permeabilized and stained with anti-cardiac troponin T (Alexa Fluor 647) and DAPI for viability and identity.

Cardiomyocytes (CM) integrity was used as an indicator of preserved cellular viability. Samples were analyzed using a NovoCyte Penteon flow cytometer (Agilent) and gated to distinguish intact cardiomyocytes from debris^11^. Controls included no-troponin staining and fluorescence compensation beads. A detailed protocol is provided in the Supplemental Material and Methods.

### RNA Extraction and Bulk RNA Sequencing Analysis

Total RNA was extracted from flash-frozen myocardial tissue using the RNeasy Mini Kit (Qiagen) and quantified with Qubit and NanoDrop. RNA integrity was assessed with an Agilent Bioanalyzer; samples with RIN > 6 were used for library preparation. <u>First Study Arm</u>: RNA-seq libraries were generated with the Illumina TruSeq Stranded mRNA kit and sequenced as 2×75 bp paired-end reads on the Aviti platform. Raw reads underwent quality control, trimming, and alignment to the *Sus scrofa* reference genome (Sscrofa11.1) using HISAT2. Gene expression was quantified with featureCounts and analyzed for differential expression with DESeq2. Genes with adjusted *p* < 0.05 and |log2FC| > 0.5 were considered significant. <u>Second Study Arm</u>: RNA-seq libraries were generated and sequenced as 2×150 bp paired-end reads by Azenta Life Sciences (Burlington, MA, USA). Sequence reads were trimmed to remove possible adapter sequences and nucleotides with poor quality using Trimmomatic v.0.36. The trimmed reads were mapped to the Pig_ERCC reference genome available on ENSEMBL using the STAR aligner v.2.5.2b. BAM files were generated as a result of this step. Unique gene hit counts were calculated by using featureCounts from the Subread package v.1.5.2. A comparison of gene expression between the groups of samples was performed using DESeq2. The Wald test was used to generate p-values and log2 fold changes. Genes with an adjusted p-value < 0.05 and |log2FC| > 1 were considered differentially expressed genes for each comparison and were visualized in volcano plots. Heatmaps were generated using the DataMap (v0.11)^12^.

The analysis of GO enrichment and KEGG pathways was performed using ShinyGO (V0.82), and visualizations were generated in ggplot2. A detailed protocol is provided in the Supplemental Material and Methods.*Whole Untargeted Metabolomics.* Global metabolomic analysis was performed on flash-frozen left ventricular biopsies collected at multiple time points (up to 48 hours) from hearts preserved with either HOPE or SCS. Tissue homogenates were spiked with isotope-labeled internal standards and extracted using chilled methanol-acetonitrile precipitation. Samples were analyzed by ultrahigh-performance liquid chromatography–high-resolution accurate mass spectrometry (UHPLC-HRAM; Thermo Exploris 240)^13,14^. Both hydrophilic interaction liquid chromatography (HILIC) and reverse-phase (C18) separation were performed under positive and negative ion modes. Spectral data were processed using Compound Discoverer 3.3 for metabolite identification and quantification. Differential analysis was conducted using fold change, p-values, and q-values, with results classified as either whole feature lists or annotated metabolite lists. A detailed protocol is provided in the Supplemental Material and Methods.

### Statistical analysis

Data are shown as mean ± standard deviation/standard error. A two-way ANOVA test, followed by a Tukey or Sidak post hoc analysis, was used to test heart weight differences between groups at different times. The Mann-Whitney test was used to assess the statistical significance of differential cardiomyocyte integrity between groups in the first study arm, and between HOPE 24h and SCS 24h post-NMP. The Kruskal-Wallis test was used to test differences between HOPE 3h, 24h, and 48h post-NMP. EPOC analysis of the perfusate was performed using the Kruskal-Wallis test. Differences were considered statistically significant when the p-value was < 0.05. Data analyses and graphical presentations were performed with GraphPad Prism 7 software.

## Results

### Study Design

The experimental design consisted of two arms (**Supplemental Fig. 1**). The first arm, Panel A, focused on myocardial edema, histological analysis, cardiomyocyte integrity, and omics of extended preservation with either SCS or HOPE. While early time points enable direct comparison, later time points extend beyond the clinically acceptable SCS window. Thus, we can evaluate the capacity of HOPE to sustain cardiomyocyte viability, transcriptional and metabolomics integrity over prolonged *ex vivo* preservation conditions not yet tested clinically. Endomyocardial biopsies were collected from ten donor hearts, SCS (n=5) and HOPE (n=5), at 0, 12, 24, and 48 hours for histology, flow cytometry, RNA sequencing, and untargeted metabolomics. In the second arm, Panel B, we employed a bench-top normothermic reanimation model (**Supplemental Fig. 2)** to assess sinus rhythm recovery and post-reperfusion cardiomyocyte integrity following extended storage. In this modified functional-recovery arm, hearts preserved with HOPE for 3h (n=2) served as a positive control, reflecting current clinical practice. Additionally, HOPE grafts were stored for 24h and 48h (n=2 each), 6–8 times longer than the routine clinical limits.

### Histopathological analysis of porcine hearts preserved with either static cold storage or HOPE

Ten cardiectomies were performed, with five hearts preserved under SCS conditions and five hearts preserved using HOPE for up to 48 hours (**Fig.1A**), a timepoint never tested before in any clinical or preclinical studies. Baseline heart weight, defined for both groups as the time before preservation commenced, was similar between SCS and HOPE organs, measuring 204.33 ± 3.51 g and 208.4 ± 13.5 g, respectively. Following 48 hours of preservation, SCS hearts experienced a 1.9% weight gain (208.33 ± 19 g) compared to baseline, while hearts preserved with HOPE demonstrated a significant 42.1% increase in weight from baseline (294.8 ± 26.99 g) (**Fig.1B**). Gross pathological evaluation of the heart did not reveal any major differences between SCS and HOPE hearts at baseline (**Supplemental Fig.3**). Histological analysis with H&E and trichrome staining of the RV septum revealed significant histopathological changes over time in either group. Edema accumulation was observed with a temporal trend throughout HOPE heart biopsies by H&E staining, but not in SCS. H&E staining of tissue of HOPE at 48h revealed few focal areas of cardiomyocyte necrosis (**Fig.1C**). No perivascular inflammation or interstitial fibrosis was observed in either HOPE or SCS **at all time points as shown** by trichrome staining (**Fig.1C**, and **Supplemental Fig.4A**). Similar results were also observed in left ventricle tissues stained both with H&E and trichrome (**Supplemental Fig.4B**). For hearts preserved with HOPE, we analyzed the perfusate via blood gas analysis at baseline and at each time point. The analysis revealed no significant differences in blood gas, electrolyte, and metabolite results in the perfusate between baseline and other time points (**Supplemental Table 1**).

**Figure 1.**
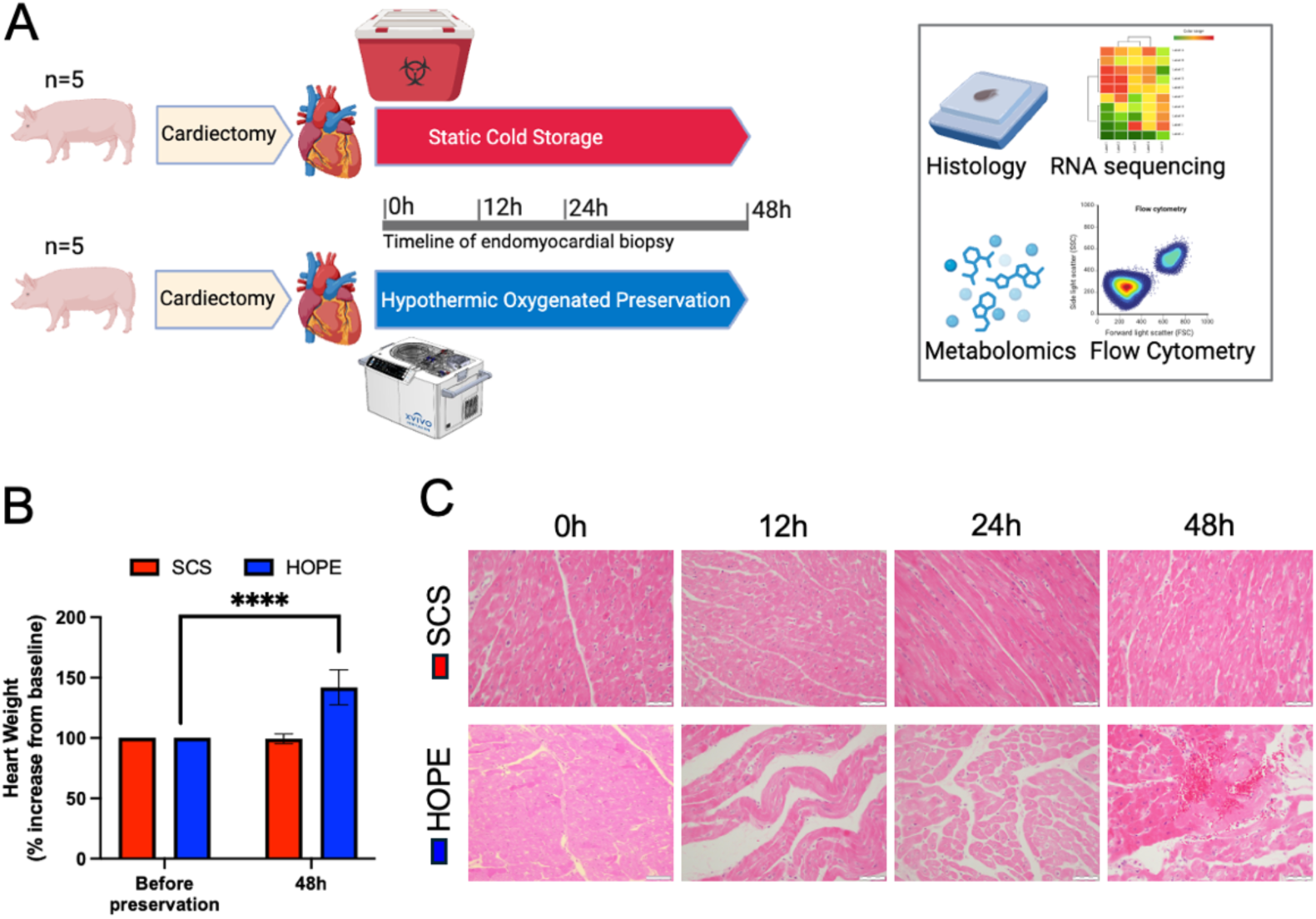
Gross and Histopathological Assessment of Cardiac Tissue Edema. **A)** Ten cardiectomies were performed in female Yorkshire pigs as described in the Methods, with five hearts preserved under SCS conditions and five hearts preserved using HOPE for up to 48 hours. Endomyocardial biopsies were collected at multiple time points (0h, 12h, 24h, and 48h) for histology, flow cytometry, RNA sequencing, and untargeted metabolomic profiling; **B)** Comparison of percent increase in heart weight following 48h SCS (red) vs HOPE (blue) preservation (n = 5 per group). Data represents the average ± SD (n=5 per group). Statistical significance was assessed using two-way ANOVA and Turkey’s post hoc analysis (**** p < 0.0001). **C)** Representative images of H&E analysis for RV septum biopsies from SCS and HOPE hearts at various time-points (0h, 12h, 24h, 48h). Formalin-fixed tissues. Scale bar = 50µm.

### Extended hypothermic oxygenated preservation conditions preserve cardiomyocyte integrity when compared to static cold storage

To examine the effects of hypothermic oxygenated perfusion on cell viability, we isolated cells from myocardial biopsies at different time points (0h, 12h, 24h, and 48h) from both SCS and HOPE hearts. Biopsies were enzymatically digested, and tissue dissociated to single-cell solution. Cells were analyzed by flow cytometry and nucleated cells were identified by DAPI staining. Among the nucleated, CM were identified by first gating using anti-cardiac troponin T. All troponin-positive events were then gated by SSC-A and FSC-A to isolate structurally intact cells from troponin-positive debris or small apoptotic bodies. FSC and SSC gating is a crucial parameter to differentiate between intact CM and small fragmented cells or debris, reducing the chance of false positives (**Fig. 2A).** Cardiomyocyte integrity was assessed by evaluating cell size and troponin expression, serving as indicators of preserved cellular functionality and viability. Analysis of the gated CM population showed a significant difference in percentage of CM when comparing SCS and HOPE at 24h and 48h of preservation (**Fig. 2B**). While identical CM were recovered at baseline, the number of cells dramatically decreased in SCS over time, with virtually no alive CM recovered at the 24h or 48h timepoints. On the contrary, CM were preserved up to 48h using HOPE.

**Figure 2.**
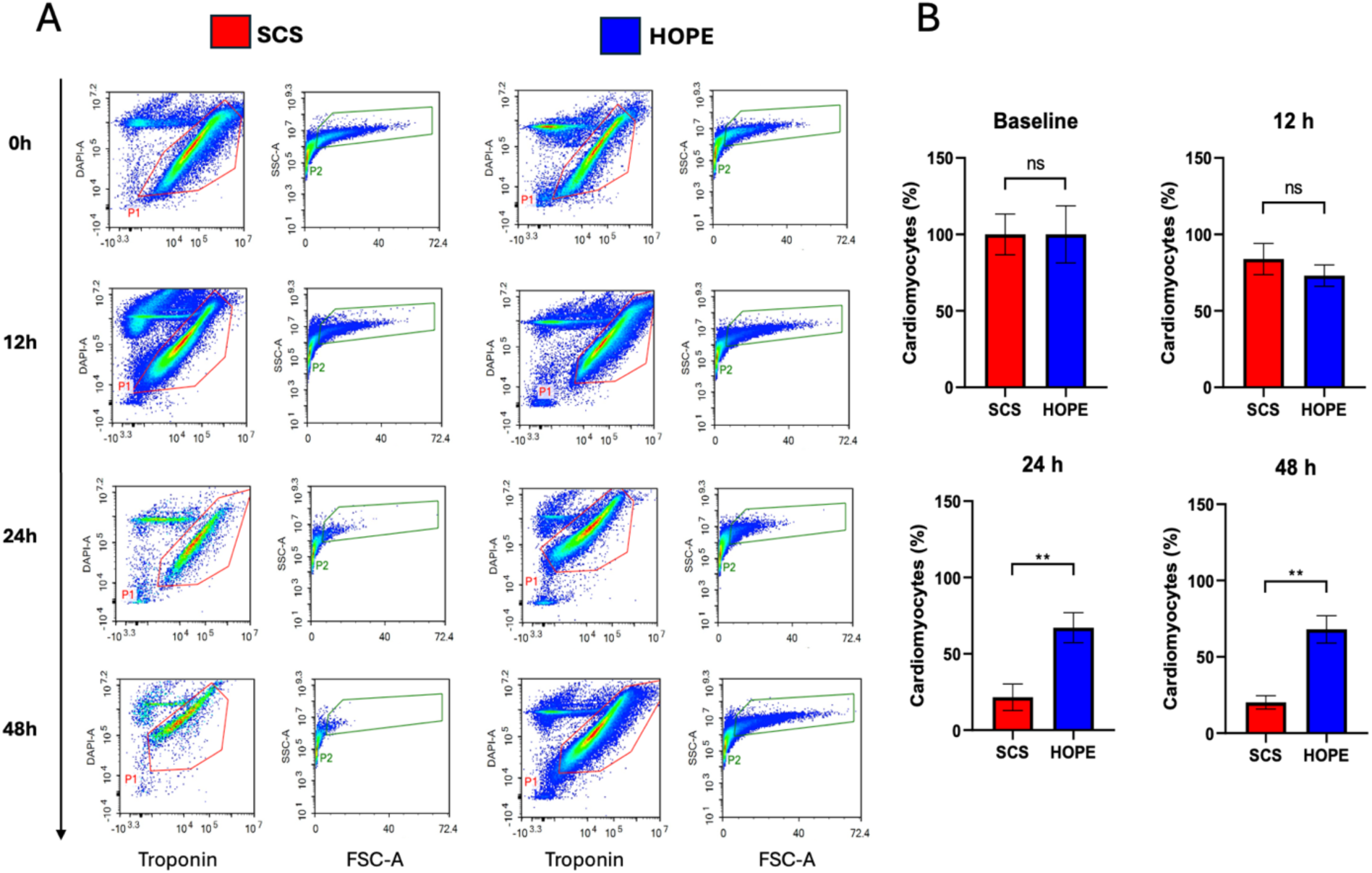
Isolation of intact cardiomyocytes in SCS vs HOPE. (A) Representative flow cytometry plots at 0h, 12h, 24h, and 48h comparing SCS and HOPE. Cardiomyocytes were gated as DAPI-negative, cardiac troponin T–positive events (P1), with intact cells further selected based on FSC/SSC properties (P2). While both preservation methods maintain cardiomyocyte viability at early time points (0–12h), viability markedly declines in SCS by 24h and 48h, with preserved gating profiles only observed in HOPE samples. (B) Quantification of viable cardiomyocytes across time points. At baseline and 12h, cardiomyocyte yields are similar between groups. However, by 24h and 48h, HOPE-preserved hearts retain significantly more viable cardiomyocytes compared to SCS. Data are presented as mean ± SEM (n = 2 per group). Statistical significance was assessed using the Mann-Whitney U test (** p < 0.01).

### RNA-sequencing analysis: an overtime comparison between SCS and HOPE heart tissue reveals differentially expressed genes related to metabolic pathways, apoptosis, and mitochondrial translation

#### Differentially expressed gene (DEG) analysis

We performed RNA sequencing and Gene Ontology (GO) and Kyoto Encyclopedia of Genes and Genomes (KEGG) pathway analysis on myocardial tissue collected at 0h, 12h, 24h, and 48h from hearts preserved with either SCS or HOPE. RNA was extracted from RV septum myocardial biopsies. The RNA integrity of SCS hearts showed consistent degradation over time, while it was preserved in all analyzed biopsies collected after HOPE; we observed that samples from the same pig are clustered in the UMAP plot (**Supplemental Fig. 5**). As expected, our DEG analysis identified no differences at baseline; we then observed a progressive increase in detected DEGs when comparing HOPE and SCS at each following time point: 132 DEGs were identified after 12h, a number that raised to 1644 DEGs at 24h and 1940 DEGs at 48h (**Fig. 3A**) (padj <0.05 & |log2FC| >0.5). The complete sets of data have been deposited and are accessible at GEO #[<u>LINK</u>]. A volcano plot and a heatmap were generated to display the distribution of DEGs and highlight the similarities of the two preservation systems at earlier time points (0h to 12h) and their divergence at later time points (24h and 48h). Bulk RNA-seq identified minimal gene-expression differences at baseline and 12h, but striking divergence at later time points. Baseline and 12-h comparisons (**Fig. B-C**) display sparse, low-magnitude changes, consistent with the expectation that early SCS and HOPE hearts remain transcriptionally similar. By 24-h, HOPE hearts up-regulate metabolic and mitochondrial transcripts (e.g., SERPINE2, NAP1L5, CKD5RAP1) and down-regulate stress-response genes (**Fig. 3D**). The divergence intensifies at 48 h (**Fig. 3E**), with HOPE showing broad up-regulation of oxidative-metabolism genes (ND4, COX1/3, CYTB) and suppression of apoptotic and inflammatory mediators compared with SCS. Heat-maps beneath each volcano plot illustrate these gene sets clustering more distinctly by preservation method at 24-h and 48-h, whereas 0 -h and 12-h samples demonstrate less variation in gene expression profile. Other key genes involved ROS, glycation, mitochondrial activity-related pathways, and apoptosis, are listed in **Supplemental Table 2**. Collectively, the data indicate that HOPE maintains a pro-metabolic, cardioprotective transcriptional profile during extended preservation, while SCS hearts have demonstrated both injury and death pathway activation beyond the 6-hour current clinical standard.

**Figure 3.**
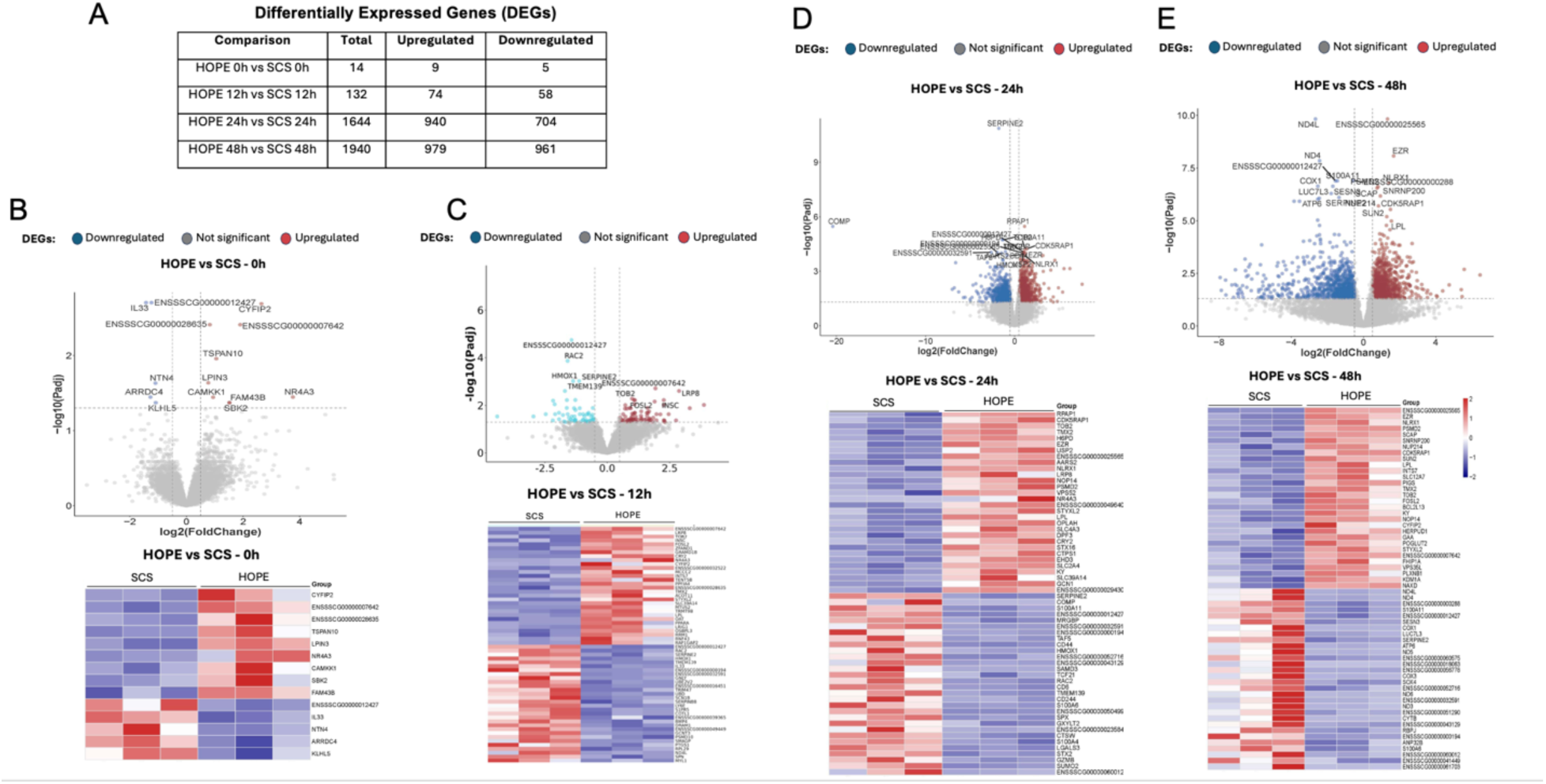
Transcriptomic differences between HOPE-and SCS-preserved hearts emerge during extended storage. (A) Summary table of differentially expressed genes (DEGs) between HOPE and SCS at 0h, 12h, 24h, and 48h, based on RNA sequencing. (B–C) Volcano plots and heatmaps for the 0h and 12h comparisons confirm the relative similarity between groups, with only a small number of genes meeting the significance threshold for differential expression (padj < 0.05, |log2FC| > 0.5). (D–E) At 24h and 48h, however, large transcriptomic shifts emerge, with HOPE-preserved hearts showing substantial upregulation of genes associated with mitochondrial activity, oxidative stress response, and anti-apoptotic signaling, while SCS hearts exhibit downregulation of metabolic and survival pathways. Heatmaps show clear clustering of HOPE and SCS samples at 24h and 48h, demonstrating progressive divergence in gene expression patterns.

#### Functional and pathway analysis

We further interrogated the complete significant DEG list of the bulk RNA-seq data to investigate the cellular and molecular functions of the genes associated with organ integrity and their related pathways. GO analysis of the DEGs revealed that at 24 hours in HOPE there was a predominant upregulation of genes involved in several metabolic pathways, such as the TCA cycle and cell metabolic processes. Further, the adrenergic signaling in cardiomyocytes, mitochondrial translation, and gene expression were upregulated in HOPE. At 48h, along with these metabolic pathways, we also observed upregulation of pathways related to longevity regulating, such as superoxide-dismutase (SOD), and adrenergic signaling in cardiomyocytes (**Fig. 4A**). Interestingly, the KEGG pathway analysis of HOPE at 24h demonstrated downregulated DEG within biological processes, including pathways associated with apoptosis, fluid shear stress, atherosclerosis, and NF-kappa B inflammatory signaling pathway (**Fig. 4B**; **Supplemental Fig. 6**). Furthermore, we noted a significant downregulation of pathways related to immune and defense responses in the HOPE tissue. Of note, only at HOPE 48h we observed fewer GO pathways associated with downregulated genes and with FDRs >1*10^5^ (reliable cutoff); some genes downregulated were associated with ribosome function and carcinogenesis-ROS-related pathways, potentially indicating a decrease in mitochondrial function. Taken together, pathway analysis demonstrates that HOPE sustains energy metabolism and cardioprotective signaling while attenuating inflammation and apoptosis during prolonged preservation.

**Figure 4.**
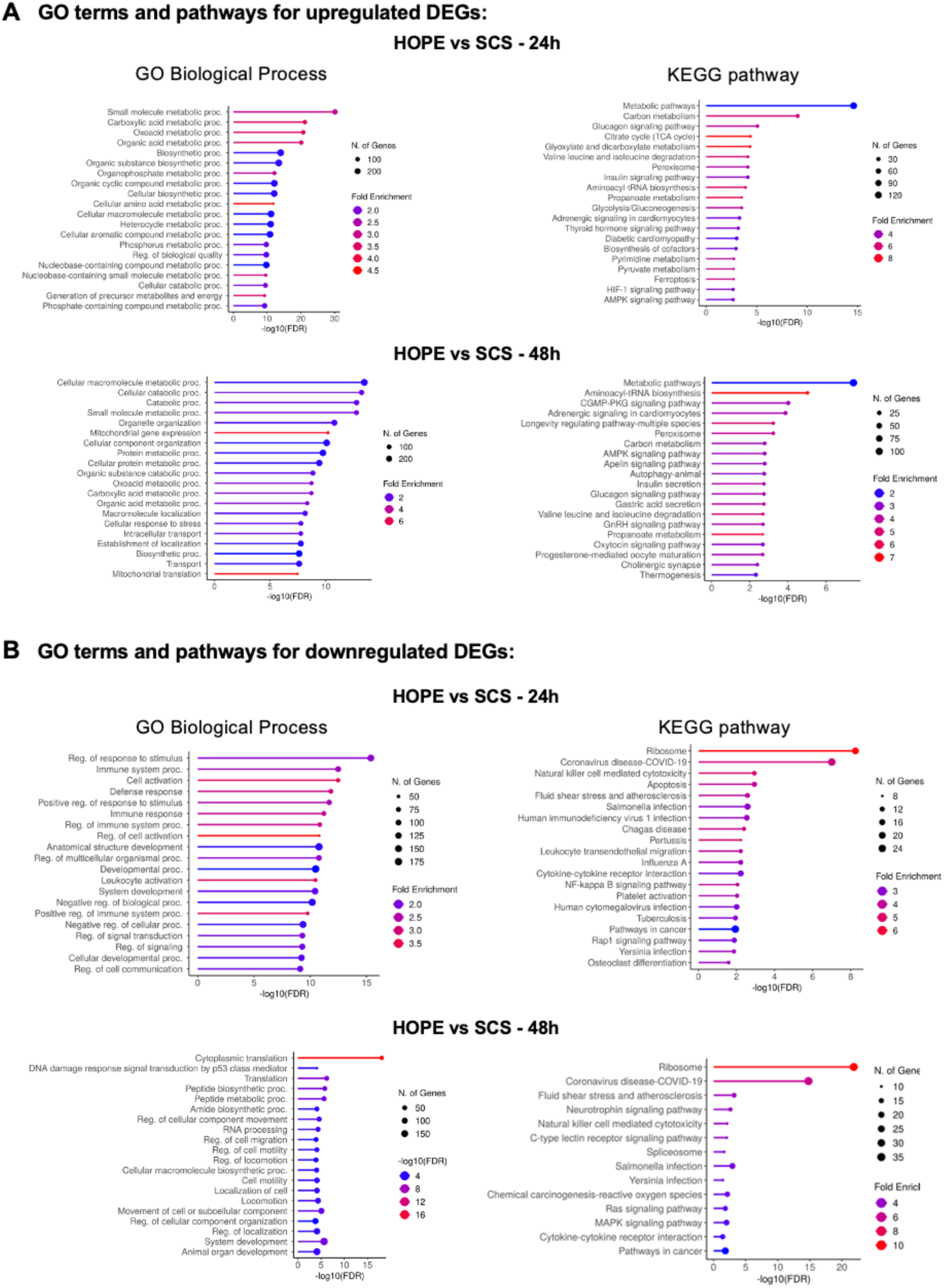
Gene ontology (GO) and KEGG pathway enrichment analyses of differentially expressed genes in HOPE vs. SCS at 24h and 48h. **(A**) Enrichment analysis of upregulated genes in HOPE-preserved hearts compared to SCS at 24h (top) and 48h (bottom). GO Biological Process analysis (left) shows significant upregulation of metabolic and biosynthetic pathways, including small molecule metabolism, mitochondrial organization, and protein catabolism. KEGG pathway analysis (right) reveals overrepresentation of energy metabolism, oxidative phosphorylation, TCA cycle, and cardiomyocyte signaling pathways such as adrenergic and AMPK signaling. These pathways reflect preserved mitochondrial function and metabolic adaptability under prolonged HOPE conditions. (**B**) Enrichment analysis of downregulated genes in HOPE vs. SCS at 24h and 48h. GO Biological Process terms (left) show downregulation of inflammatory and immune response pathways, including cytokine signaling, leukocyte activation, and stress-response regulation. KEGG pathway maps (right) reinforce this pattern, showing reduced enrichment of apoptosis, fluid shear stress, and infection-related signaling in HOPE-preserved hearts.

### Metabolomics analysis of endomyocardial biopsies obtained from porcine hearts preserved with either SCS or HOPE

We performed whole untargeted metabolomics on the LV endomyocardial biopsies to ascertain differences in metabolite expression between the two groups. The complete sets of data have been deposited and are accessible at GEO #[<u>LINK</u>]. Untargeted metabolomic profiling (BCL)-condensed analysis, represented with a PCA plot, showed strong segregation between SCS and HOPE sample groups, both among whole features and annotated compounds (**Fig. 5A**). The analysis identified 5456 whole features and 651 annotated compounds, suggesting the whole metabolomic profile of the annotated feature list was representative of the whole feature list. Heatmaps of the whole feature (**Fig.5B top**) and of the annotated compound list (**Fig.5B bottom**) showed distinct and replicable differential metabolomic profiles in the SCS versus HOPE preservation groups when clustered by time point and pig group (FDR <0.05 and |log2 fold change|>0.693). Volcano plots of the annotated compounds revealed the distribution of the differentially expressed metabolites and highlighted the top upregulated and downregulated compounds at each time point (**Fig.5C**). Of note, the differential metabolomic profile of SCS vs. HOPE hearts increased over 48 hours of storage.

**Figure 5.**
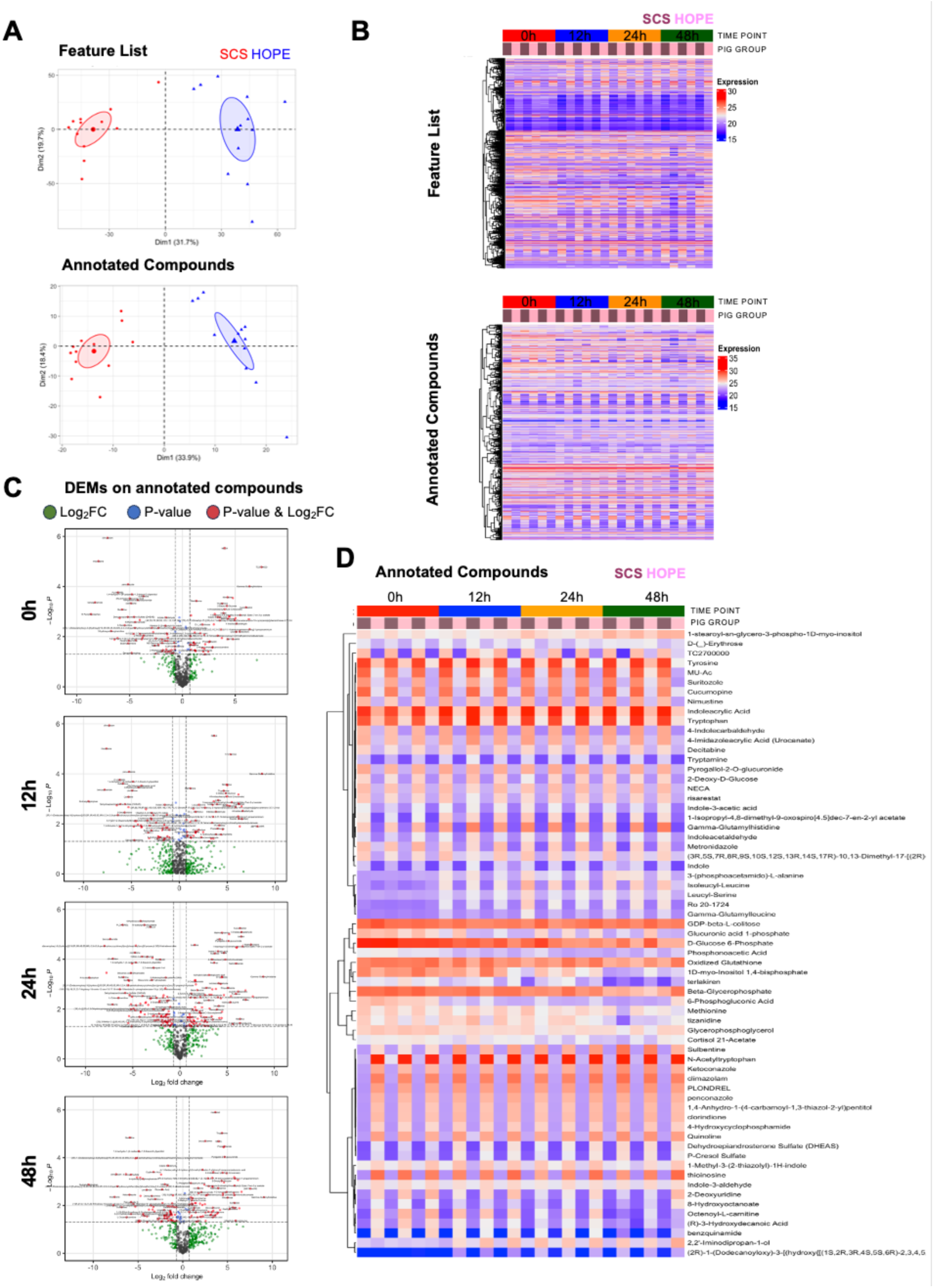
Metabolomic analysis for HOPE vs SCS. **A**) Unsupervised dimensionality reduction of whole feature list and annotated compounds showing segregation of SCS (red) and HOPE (blue) pigs. **B)** Supervised clustering of whole feature list and annotated compounds showing similar clustering by SCS (brown) and HOPE (pink) and by time point (0h, 12h, 24h, 48h) (padj <0.05 & abs(log2FC) >0.5). **C)** Volcano plots showing increasing divergence between whole metabolomic profile of SCS vs HOPE with time (0h to 48h). **D)** Supervised clustering of top 65 annotated compound metabolites shows similar clustering by SCS (brown) and HOPE (pink) and by timepoint (0h, 12h, 24h, 48h) (padj <0.05 & abs(log2FC) >0.5).

Furthermore, functional analysis of the top 65 differentially expressed metabolites showed greater accumulation of key metabolites in the SCS groups, including but not limited to reduced glutathione (GSH), Xanthine and Hypoxanthine metabolites, and lower accumulation of arachidonic acid and eicosanoids, all suggestive of greater ischemic/reperfusion injury of the myocardium in SCS groups (**Fig. 5D**). Further, we performed pathway-based functional analysis of SCS versus HOPE samples at various time points to see differences in metabolite levels between compounds belonging to known metabolic pathways (**Supplemental Table 3**). We compared key metabolites in the TCA cycle, glycolysis, and ROS-related pathways across 48h HOPE and SCS samples. Notably, AMP was significantly elevated in SCS vs. HOPE at 48h (p=0.008), consistent with purine breakdown and energy failure in ischemic tissue. In contrast, FAD (flavin adenine dinucleotide) levels were significantly reduced in SCS relative to HOPE (p= 0.018), suggesting impaired mitochondrial cofactor availability. Markers of oxidative stress diverged strongly between groups. Arachidonic acid, a lipid peroxidation substrate, was significantly depleted in SCS vs HOPE (p= 0.008), indicating higher turnover or damage in SCS. Oxidized glutathione (GSSG) was also lower in SCS (p= 0.073) and dropped dramatically in SCS compared to baseline (p= 0.0008), pointing to loss of antioxidant capacity. In contrast, reduced glutathione remained relatively stable in HOPE vs baseline (ns), whereas it declined significantly in SCS (p= 0.024). Together, these findings suggest that HOPE preserves metabolic cofactors and antioxidant defenses during prolonged preservation, while SCS is associated with energetic collapse and redox imbalance by 48 hours. These metabolic data support the concept that HOPE hearts maintain mitochondrial energy flux, limit oxidative damage and thereby preserve contractile competence well beyond the 6-h cold-storage threshold.

### Functional recovery of HOPE-preserved hearts following bench-top normothermic reperfusion

In a second set of experiments, 6 hearts were procured and preserved with HOPE for 3h, 24h, and 48h (n=2/group) conditions, followed by a 2-hour period of Normothermic Machine Perfusion (NMP), (**Fig. 6A**). A single heart kept in SCS for 24h (n=1) was included as a negative control, as decades of experience indicate that such prolonged cold storage cannot support reanimation and was deemed ethically unnecessary. The 2h reperfusion time frame was chosen to replicate the early reperfusion period after clinical implantation, during which myocardial function, rhythm recovery, and viability can be reliably evaluated.

**Figure 6.**
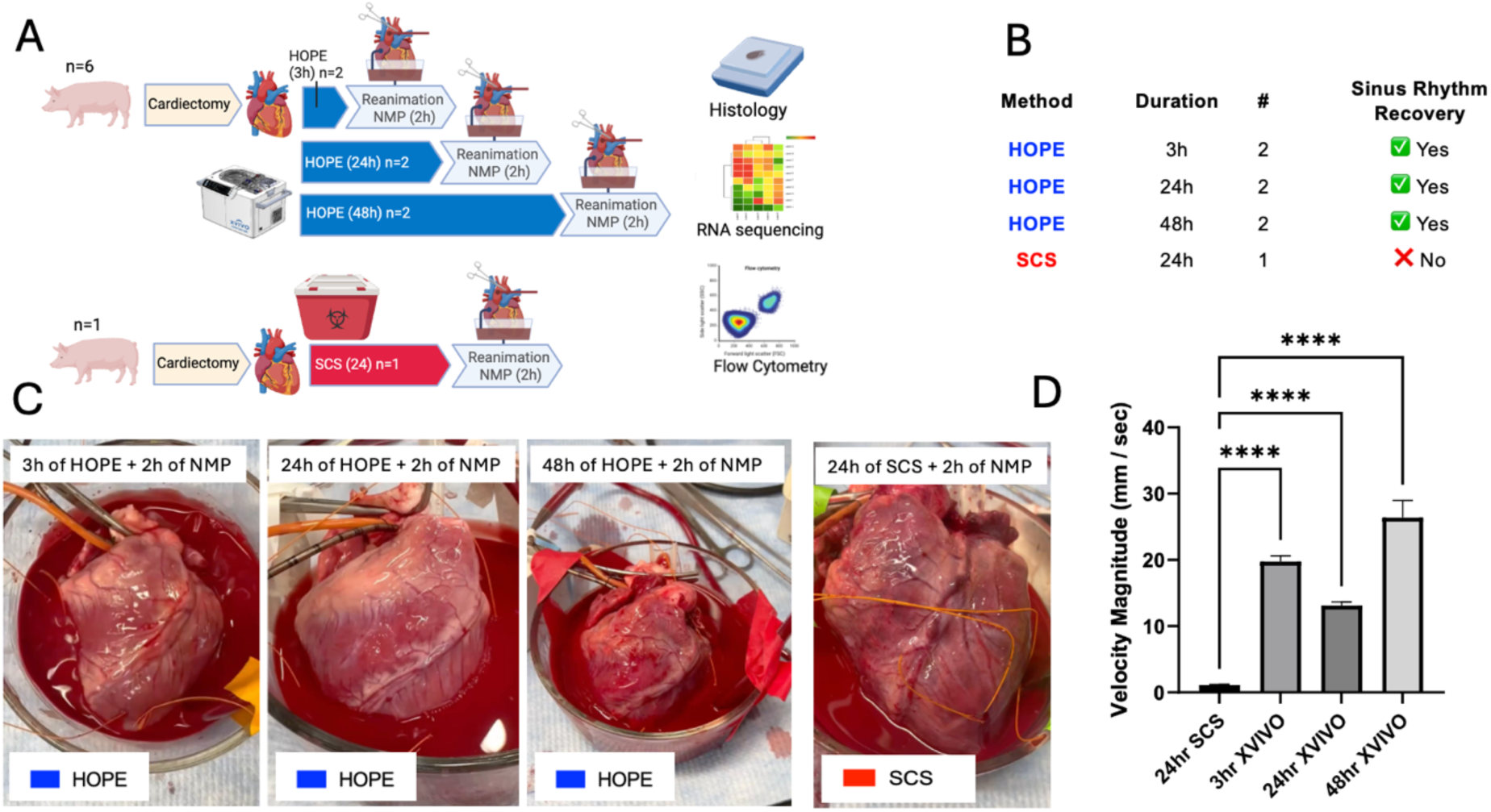
Functional recovery following reanimation of HOPE-and SCS-preserved hearts. (A) Schematic experimental design for the functional recovery arm (Arm 2). Porcine donor hearts were preserved using hypothermic oxygenated perfusion (HOPE) for 3 h (n = 2), 24h (n = 2), or 48h (n = 2), or static cold storage (SCS) for 24h (n = 1), followed by 2 hours of normothermic machine perfusion (NMP) to simulate transplant reperfusion. After reanimation, hearts were assessed for rhythm recovery, gross morphology, cardiomyocyte viability via flow cytometry, and bulk-RNA sequencing analysis. (B) Summary table showing that all HOPE-preserved hearts regained normal sinus rhythm, including those stored for up to 48h. In contrast, the SCS-preserved heart failed to reanimate. (C) Representative still frames from video recordings after 2h of NMP. HOPE hearts at 3h, 24h, and 48h exhibited robust, coordinated contraction, while the SCS heart showed no organized motion. (HOPE 3h + 2h NMP, **Supplemental Video 1:** DOI <u>Link</u> - HOPE 24h + 2h NMP **Supplemental Video 2** DOI <u>Link</u> - HOPE 48h + 2h NMP **Supplemental Video 3** DOI <u>Link</u> - SCS 24h + 2h NMP **Supplemental Video 4** DOI: <u>Link</u>); (D) Average velocity magnitude (mm/s) ± SEM of each group at all analyzed timepoints. ****p<0.0001 by one-way ANOVA with Dunnett’s multiple comparison test.

As anticipated, the 3h HOPE heart readily resumed normal sinus rhythm during the 2h of normothermic NMP reperfusion (**Supplemental Video 1:** HOPE 3h + 2hNMP: DOI: <u>Link</u>) remarkably, hearts preserved for the far longer intervals of 24-h and 48-h also re-established stable sinus rhythm (**Fig.6B-C**; **Supplemental Video 2** HOPE 24h + 2hNMP: DOI: <u>Link</u>; **Supplemental Video 3** HOPE 48h + 2hNMP: DOI: <u>Link</u>). In contrast, the 24 h SCS heart failed to regain rhythm under identical NMP conditions (**Supplemental Video 4**: SCS 24h + 2h NMP: DOI: <u>Link</u>). The optical flow analysis of frame-to-frame displacement confirmed that significant contractile activity was regained in HOPE storage hearts, with significantly elevated average velocity magnitudes across all contraction cycles at timepoints 3h, 24h, and 48h. Conversely, the recovery of contractility on an NMP circuit was absent in a 24h SCS heart (**Fig. 6D, Supplemental Fig. 7**). The data suggest that HOPE may enable recovery of hearts stored for up to 48 hours, as evidenced by benchtop reanimation of contractility and observed sinus rhythm following normothermic reperfusion. In contrast, static cold storage for 24 hours fails to support electrical reanimation, highlighting the superior capacity of HOPE to maintain myocardial viability and electrophysiological integrity during extended *ex vivo* preservation.

### Gross and Histologic Pathology Reveals a Preservation Limit Between 24 h and 48 h for HOPE

For all these 7 hearts, weight was recorded before preservation, after preservation with HOPE 3h, 24, 48h and after 2h of NMP. Organ weight increased following 3h, 24h, and 48h of HOPE preservation, with more weight gain noted with increasing preservation time (3h HOPE: 114.05 ± 1.7%, 24h HOPE: 142.49 ± 3.5%, 48h HOPE: 150 ± 10.73%). Hearts that underwent 3h or 24h HOPE preservation did not exhibit a significant increase in weight after NMP. However, HOPE hearts preserved for 48h continued to see a significant change in weight gain after reperfusion of 189.84 ± 5% increase from baseline (**Fig. 7A**).

**Figure 7.**
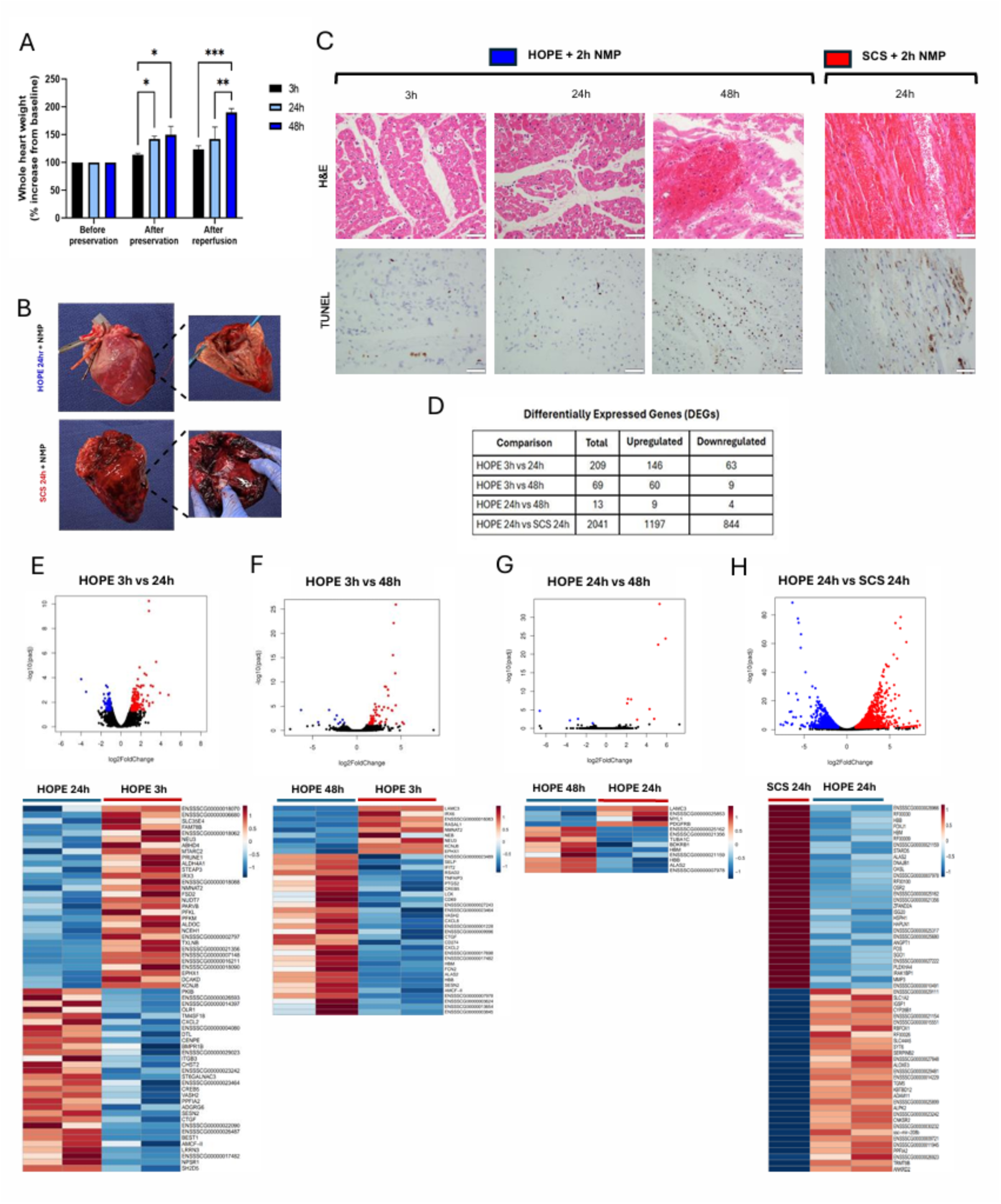
Gross and Histologic Pathology Reveals a Preservation Limit Between 24h and 48h for HOPE. (**A**) Quantification of heart weight (% increase from baseline) before preservation, after preservation, and after 2-hour NMP. Weight gain was progressive with longer HOPE durations and increased further after reperfusion at 48h. Data represent mean ± SD; *p<0.05, **p<0.01, ***p<0.001 by two-way ANOVA with Tukey’s post hoc test. (**B**) Gross pathology following NMP. HOPE hearts exhibited preserved morphology, while the SCS 24h heart showed diffuse hemorrhage and transmural necrosis. (**C**) Representative histologic sections of left ventricular myocardium after NMP. H&E staining shows preserved architecture in HOPE 3h and 24h, with edema and focal necrosis in HOPE 48h and severe injury in SCS 24h. TUNEL staining highlights limited apoptosis in HOPE groups and extensive cardiomyocyte apoptosis in SCS. Scale bars = 50 µm. (**D**) Summary table of differentially expressed genes (DEGs) between HOPE at 3h, 24h and 48h and SCS at 24h followed by 2h NMP, based on RNA sequencing. (**E-G**) Volcano plots and heatmaps for the HOPE timepoints comparisons confirm the relative similarity between groups, with only a small number of genes meeting the significance threshold for differential expression (padj < 0.05, |log2FC| > 1). (**H**) Volcano plot and heatmap for the HOPE 24h vs SCS 24h + NMP 2h comparison show a large divergence in transcriptomic patterns.

Gross examination of HOPE hearts preserved for 48 hours followed by NMP showed scattered petechiae along with focal areas of myocardial necrosis. These findings were not present in HOPE hearts preserved for 3 or 24 hours followed by reperfusion (**Fig. 7B**), indicating that, under the current protocol, the threshold for optimal preservation may fall between 24 and 48 hours. In the heart preserved under SCS conditions for 24 hours and reperfused with NMP, gross pathology after reperfusion revealed large transmural segments of myocardial necrosis (**Fig. 7B**). These findings are consistent with those shown in **Figs. 1**-**5** and align with existing literature highlighting the limitations of prolonged static cold storage over a 3-6 hour limit.

H&E staining of HOPE hearts reanimated with NMP showed interstitial widening consistent with edema. Coagulative myocyte necrosis was noted to be present in the left ventricular free wall, but only in HOPE hearts preserved for 48 hours followed by NMP (**Fig. 7C**), potentially indicating more significant cardiomyocyte injury with time. Focal inflammation with neutrophilic infiltration was seen in one heart that was reperfused following 48 hours of preservation with HOPE. There was no evidence of interstitial fibrosis or thrombosis in these experiments, assessed by trichrome staining (**Supplemental Fig. 8**). In 3h and 24h HOPE hearts, TUNEL staining revealed sparse apoptosis of interstitial cells (i.e. interstitial fibroblasts or macrophages, inflammatory cells, and/or endothelial cells). Apoptosis of cardiomyocytes was only observed in HOPE hearts preserved for 48 hours; however, it was minimal as compared to 24hr SCS preserved hearts (**Fig. 7C**).

### RNA-sequencing analysis: a comparison of gene expression profiles between heart tissues after 3h, 24h and 48h of HOPE, and between HOPE and SCS at 24h, followed by 2h of NMP

#### Differentially expressed gene (DEG) analysis

Bulk RNA sequencing analysis was performed on RV septum myocardial biopsies collected after 3h, 24h, and 48h of HOPE preservation followed by 2h of NMP, and after 24h of SCS followed by 2h of NMP. The total number of identified DEGs in the comparisons between HOPE at 3h and 24h, HOPE at 3h and 48h, and HOPE at 24h and 48h were 209, 69 and 13, respectively. In contrast, the number of DEGs notably increased to 2041 when comparing HOPE and SCS preservation conditions at the 24h time point (**Fig. 7D**) (padj <0.05 & |log2FC| >1). The complete sets of data have been deposited and are accessible at GEO #[<u>LINK</u>]. A volcano plot and a heatmap were generated to display the distribution of DEGs. Limited transcriptomic differences emerged from the comparisons between different HOPE timepoints, with minimal changes reported between 24h and 48h of HOPE preservation. This result proved that the HOPE preservation of heart tissues maintained stable gene expression profiles over time. (**Fig.7 E-G**). A significant divergence in gene expression profiles emerged between hearts preserved in either HOPE or SCS for 24h as illustrated by the highly distinct clustering of DEGs in Volcano plot and heatmap (**Fig. 7H**).

#### Functional and pathway analysis

GO analysis of the DEGs revealed minimal to no significant pathways above the reliable FDR cutoff (FDR <0.001 for lists with fewer than 100 genes and FDR < 10E-5 for more than 100 genes, using ShinyGO software), indicating background noise due to the small to medium-sized gene lists when comparing different HOPE time points for both upregulated (**Supplemental Fig. 9**) and downregulated genes (**Supplemental Fig. 10**). When comparing the 3h vs 24h sample, only a few enriched GO biological terms emerged above the cutoff, and no significantly enriched KEGG pathways were identified. However, when comparing the 3h vs 48h samples, pathways related to genes upregulated in response to stress and defense were noted, although this was based on a limited list of only 69 genes, with only a few significantly enriched KEGG pathways. In the comparison of the 24h vs 48h HOPE samples, no pathways with more than two genes were found in the enrichment analysis. In contrast, both KEGG and GO analyses comparing SCS to HOPE at 24h showed upregulated and downregulated pathways associated with various biological processes. These included pathways related to cellular respiration, aerobic respiration, metabolic pathways, mitochondrial organization, ATP production, heart and muscle contraction, and ROS. This RNA-sequencing analysis highlights the stability of gene expression in hearts preserved with HOPE over time, contrasted by a significant transcriptomic shift in hearts preserved with SCS.

### Extended hypothermic oxygenated preservation followed by a 2-hour period of Normothermic Machine Perfusion maintain cardiomyocyte integrity when compared to static cold storage

Using 3h HOPE, the current clinical standard, as the reference condition we examined whether longer hypothermic oxygenated perfusion preserves CM integrity upon normothermic reanimation. Endocardial biopsies obtained after 2 h of NMP were dissociated and analyzed by flow cytometry. CM were defined as DAPI-negative, cardiac troponin-T–positive events (P1) that fell within an intact-cell FSC/SSC gate (P2). Representative plots show robust P1/P2 populations at 3h, 24h, and 48h with HOPE (**Fig. 8A** and **B**). Quantitatively, CM yield at 24-h and 48-h did not differ significantly from 3 h (**Fig. 8C**), indicating that HOPE preserves CM integrity even after 48h, six-to eight-fold longer than routine cold storage. A single 24h SCS heart served as a negative control. Its post-NMP cytogram displayed a collapsed troponin-positive gate and virtually no intact-cell events (**Fig. 8D**); viable CM were < 5 % of the HOPE cohort (**Fig. 8E**).

**Figure 8.**
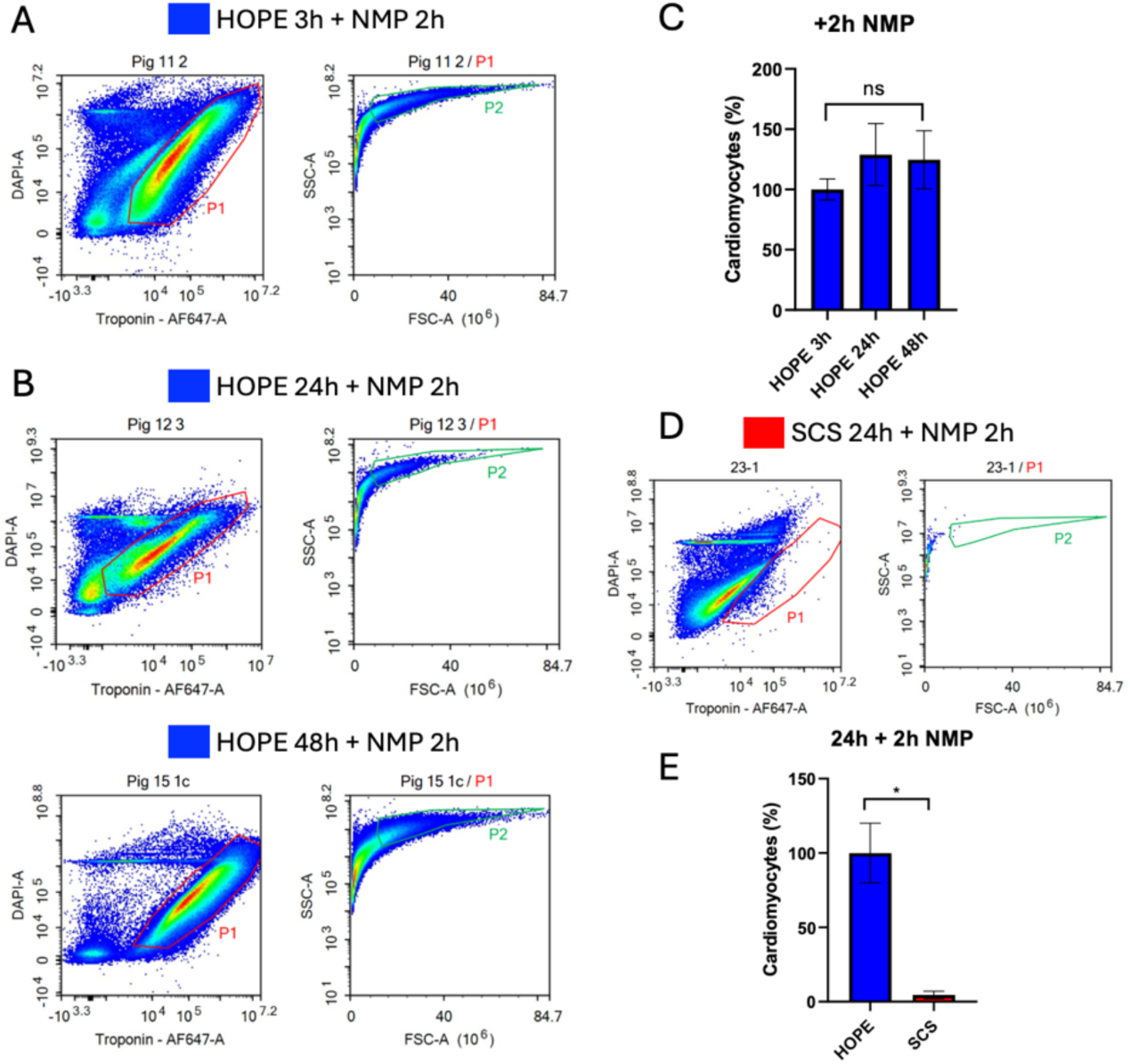
Flow cytometry quantification of intact cardiomyocytes following normothermic reperfusion. Representative flow cytometry plots of hearts preserved with hypothermic oxygenated perfusion (HOPE) for 3 h (the current clinical standard) (**A**) or 24h and 48h (**B**), followed by 2 h of normothermic machine perfusion (NMP). Cardiomyocytes were identified as DAPI-negative, cardiac troponin T–positive cells (P1), with subsequent FSC/SSC gating to isolate intact cells (P2). (**C**) Quantification of cardiomyocyte yield across all HOPE time points after NMP shows preservedintegrity and comparable recovery at 3h, 24h, and 48h (n=2 per group). Data are presented as mean ± SEM. Statistical significance was assessed using the Kruskal–Wallis test (nonparametric one-way ANOVA; ns = not significant). (**D**) Representative flow cytometry plot of a heart preserved for 24h with static cold storage (SCS) followed by 2h NMP. A marked reduction in viable cardiomyocytes is observed, with severely diminished P1 and P2 gates. (**E**) Quantification of viable cardiomyocytes in 24h HOPE versus 24 h SCS conditions following reperfusion reveals a >25-fold reduction in CM viability in SCS-preserved hearts. Data are presented as mean ± SEM. Statistical significance was assessed using the two-tailed Mann-Whitney U test (* *p < 0.05*).

## Discussion

*Ex vivo* preservation systems are rapidly evolving and are showing increasing promise to extend the durability of cardiac preservation above the limit of SCS^15^. This study aimed to evaluate molecular, structural, and cellular integrity during extended heart preservation with SCS or HOPE, and to assess functional recovery and cardiomyocyte viability following normothermic reperfusion as a transplant simulation.

In the first arm, we showed that cardiomyocyte integrity was maintained in HOPE storage up to 48h, significantly exceeding the current standard preservation window by 6 to 8 times. Increased cell size and sustained troponin signal suggest that the cardiomyocytes retained their contractile phenotype and remained viable under these extended conditions. Deterioration of CM during dissociation protocols has been similarly reported in diseased cardiac tissue from hypertrophic cardiomyopathy or heart failure patients, suggesting SCS-derived cells are likely more susceptible to damage^16^. We show that the gene expression and metabolomic profiles of HOPE and SCS stored hearts were distinct and divergent throughout preservation. RNA enrichment profiles of HOPE pathways consistently demonstrated increased expression of cardioprotective metabolic pathway-related genes and mitochondrial-related genes. It also showed downregulated processes associated with apoptosis, fluid shear stress, atherosclerosis and NF-kappa B inflammatory signaling. Genes found in our GO analysis pathways and decreased in SCS, have been described as enhancing a cardio-protective function in several studies. For instance, NLRX1^17^ and NADX^18^ are linked to the elimination of metabolic waste, mitochondrial damage, and apoptosis of epithelial cells, particularly in an oxidative stress–dependent manner. Additionally, S100A4 is associated with promoting the growth of cardiomyocytes in injured myocardium^19^, while SERPINE2 has been studied in mouse models of cardiac fibrosis^20^. Decreased HOPE genes such as CTSW, SUMO2, LPL, when upregulated, are described promoting cardiac stress, apoptosis and fatty-acid related dysfunction in heart failure studies^21–23^. Untargeted metabolomic profiling showed differential accumulation of metabolites involved in ischemic reperfusion injury, mitochondrial dysfunction, and anerobic metabolism in HOPE vs SCS hearts. While our analysis is not sufficiently powered to make conclusive claims about organelle health, our findings, when taken in conjunction with previous research done in other organ preservation, lead us to hypothesize that SCS stored hearts are more metabolically inactive, showing more anaerobic respiration, more mitochondrial dysfunction, and undergo more ROS and ischemic reperfusion injury. With mitochondrial health, for example, our study found the accumulation of Itaconate, glutamine, FAD, in HOPE-preserved hearts, suggestive of increased mitochondrial viability.

In the second arm of the study, we utilized a bench-top normothermic reperfusion (NMP) system to assess functional recovery following prolonged heart preservation. This approach parallels the on-table reanimation of heart allografts which adapts the Langendorff perfusion model to evaluate the viability of donor hearts by restoring warm, oxygenated blood flow into the coronary circulation. This well-established technique has been recently used in clinical practice by Duke’s Pediatric and Congenital Heart Center to assess the suitability of a pediatric donor heart procured under donation after circulatory death criteria, enabling safe transplantation while avoiding the ethical concerns associated with normothermic regional perfusion. This work strengthens the efficacy of on-table reanimation and offers promise for more clinical use of this technique^24^. During reperfusion on the NMP circuit, we observed that all HOPE hearts regained normal sinus rhythm regardless of the time of perfusion in the XVIVO machine. These findings align with the bulk-RNA sequencing analysis, which revealed similar transcriptomic profiles across all HOPE preservation timepoints highlighting the transcriptomic resilience of HOPE-preserved cardiac tissues. Conversely, SCS-preserved hearts failed to regain sinus rhythm, even after 2-hour NMP, and showed a divergent transcriptome compared to HOPE hearts. This data provides further mechanistic support for the adoption of HOPE. Furthermore, flow cytometry analysis post-NMP demonstrated significantly higher percentages of intact cardiomyocytes in HOPE-preserved hearts compared to SCS, supporting the histologic and functional findings.

While we observed edema and focal injury in the 48-hour HOPE group, overall preservation of rhythm and cardiomyocyte viability supports this timepoint as a potential upper limit under current perfusion conditions. Myocardium is highly susceptible to edema formation due to its dense microvascular network and elevated interstitial flow rate, especially when the intracellular mechanisms regulating cardiomyocytes and interstitial volume are disrupted^25,26^. That HOPE stored hearts accumulate edema, even within acceptable perfusion times, has long been known^27^, and this finding was corroborated by our findings, with the amount of edema linked to the preservation time. Whether edema formation is primarily related to perfusion hydrostatic pressure, perfusate oncotic pressure, or endothelial dysfunction, however, remains to be shown, and will be the subject of future investigations.

### Limitations

This study has several limitations. First, while the bench-top normothermic reperfusion (NMP) model provided valuable insights into functional recovery following extended preservation, it does not fully replicate physiologic conditions. The heart operates in an unloaded state, without native preload or afterload, which limits the assessment of myocardial contractility, distensibility, and hemodynamic performance. While sinus rhythm and CM viability offer important insights, the lack of hemodynamic measurements limits our ability to assess inotropic function or myocardial performance under load-bearing conditions. Second, the absence of an orthotopic transplantation arm prevents evaluation of long-term graft function, integration, and post-transplant remodeling under in vivo conditions. This will be a critical next step to validate 48-hour preservation as a viable clinical strategy. It should be noted that our metabolomic profiling used different solutions when arresting the heart. SCS hearts were arrested using HTK solution, whereas HOPE hearts were arrested with XVIVO heart solution. Lastly, the study was conducted in a porcine model with relatively small sample sizes in some experimental groups, particularly the SCS+NMP arm, which may constrain statistical power and generalizability. These experiments, however, set the stage for our ongoing clinical porcine transplant model studies.

### Clinical Relevance

There are several elements of potential clinical significance for this pre-clinical study. As detailed below, our research supports the usage of this device for heart preservation for up to 24 hours or more. Additional cardioprotective mechanisms or altered preservation conditions could further expand this preservation time. Extending preservation to 24 hours or more would bring us closer to elective surgery with improved coordination and optimized resource allocation. Additionally, extended preservation times could lead to enhanced focus on refining preservation technologies, such as improved perfusion systems and preservation solutions.

## Conclusion

In summary, our study demonstrates that HOPE enables superior preservation at 24-h and 48-h time points compared to SCS. Furthermore, HOPE preservation sustained structural, cellular and molecular mechanisms well beyond conventional timeframes, establishing a foundation for future translational studies and highlighting its potential to extend clinical preservation times, improve surgical logistics, and contribute to expanding the donor pool through broader geographic sharing.

## Data Availability

All data produced in the present study are available upon reasonable request to the authors

## Acknowledgments.

We gratefully acknowledge Shaheer Khalid Faruqi and Dr. Craig J. Goergen (Purdue University, Weldon School of Biomedical Engineering) for their generous guidance and technical advice on the frame-to-frame motion-analysis workflow that quantified ventricular contractile activity during normothermic reperfusion. Histology Core: Histological specimen preparation and staining for this work were performed in the Molecular Pathology Shared Resource of the Herbert Irving Comprehensive Cancer Center at Columbia University, supported by NIH grant #P30 CA013696 (National Cancer Institute). JP Sulzberger Columbia Genome Center: “This research was funded in part through the NIH/NCI Cancer Center Support Grant P30CA013696 and used the Genomics and High Throughput Screening Shared Resource.” Columbia Stem Cell Initiative Flow Cytometry core. Columbia Biomarkers Core Laboratory.

## Author Contributions

Conceptualization: GF, CC, MKM, YK, AC, KP, JBG, KT

Methodology: CC, MKM, YK, AC, AA, KP, CK, EF, SJB, TP, AR, KT, GF

Investigation: CC, MKM, YK, AC, AA, KP, CK, EF, SJB, CK, BB, TP, LP, CC, KF, MS, DA, EB, AR, MT, JBG, KT, GF

Visualization: CC, MKM, YK, GF

Funding acquisition: GF

Writing – original draft: CC, MKM, YK, AC, CK, AA, JBG, KT, GF

Writing – review & editing: CC, MKM, YK, CK, KT, JBG, GF

## Disclosure, Competing Interests

Dr. Ferrari’s Andrew Sabin Family Foundation Cardiovascular Research Laboratory received an industry-sponsored grant from XVIVO Perfusion Inc. Authors declare that they have no competing interests.

**Data and Material Availability** [GEO # LINK for RNAseq and GEO# LINK for Metabolomics]

## List of non-standard abbreviations

HOPE: Hypothermic Oxygenated Perfusion
SCS: Static Cold Storage
NMP: Normothermic Machine Perfusion

## List of Supplemental Material

- Supplemental Fig 1. Experimental Design Overview
- Supplemental Fig 2. Normothermic Machine Perfusion Circuit Setup
- Supplemental Fig 3. Gross Pathology of HOPE and SCS Hearts After Preservation
- Supplemental Fig 4. Histopathological Assessment of the Right and Left Ventricular Myocardium During Preservation
- Supplemental Fig 5. Transcriptomic Clustering of HOPE and SCS Samples by UMAP
- Supplemental Fig 6. Enriched KEGG Pathways Among DEGs in HOPE-Preserved Hearts
- Supplemental Fig 7. Mean Velocity Magnitude graphs during 2h of Normothermic Machine Perfusion following HOPE or SCS preservation.
- Supplemental Fig 8. Trichrome Staining of RV Septum Following Normothermic Reperfusion in HOPE and SCS Hearts
- Supplemental Fig 9. Gene ontology (GO) terms and KEGG pathway enrichment analysis of significantly upregulated genes in 3, 24, and 48h HOPE preservation groups.
- Supplemental Fig 10. Gene ontology (GO) terms and KEGG pathway enrichment analysis of significantly downregulated genes in 3, 24, and 48h HOPE preservation groups.
- Supplemental Table 1. HOPE Perfusate Composition Over Time
- Supplemental Table 2. Differentially Expressed Genes and Their Related Pathways
- Supplemental Table 3. Differentially Identified Metabolites and Associated Pathways
- Supplemental Video 1. Reanimation of Heart After 3h HOPE + 2h NMP
- Supplemental Video 2. Reanimation of Heart After 24h HOPE + 2h NMP
- Supplemental Video 3. Reanimation of Heart After 48h HOPE + 2h NMP
- Supplemental Video 4. Failed Reanimation of Heart After 24h SCS + 2h NMP
- Supplemental Material and Methods

---

**Supplemental Fig 1.**
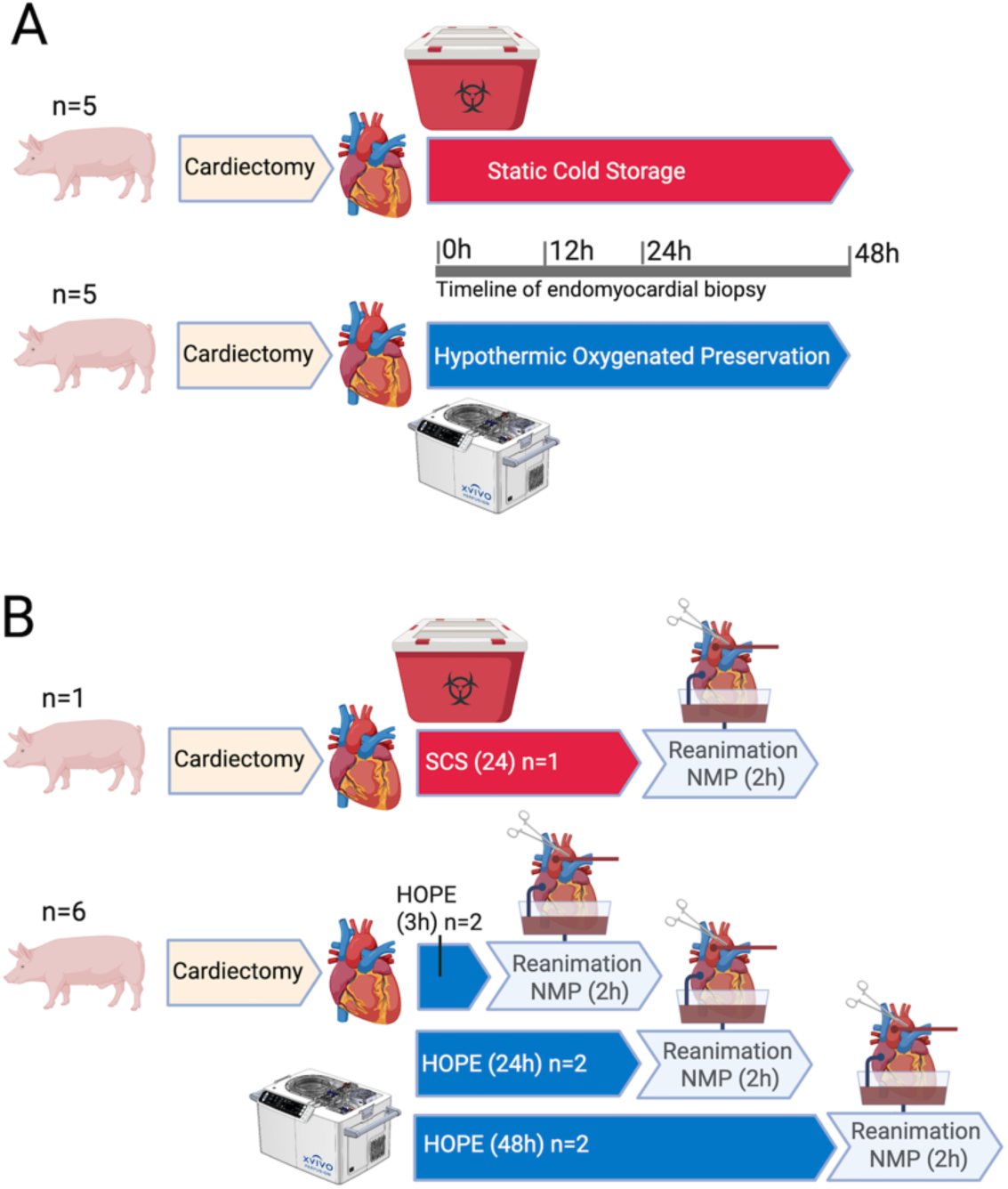
Rationale for Experimental Design. **A)** to evaluate cardiomyocyte viability, myocardial edema, histological integrity, and molecular changes during extended preservation using either static cold storage (SCS) or hypothermic oxygenated perfusion (HOPE). Endomyocardial biopsies were collected at multiple time points (0h, 12h, 24h, and 48h) for histology, flow cytometry, RNA sequencing, and untargeted metabolomic profiling; **B)** to assess the functional recovery of preserved hearts as a model of transplantation. Hearts stored for varying durations in either SCS or HOPE underwent normothermic machine perfusion (NMP) for 2 hours to simulate transplantation and test their ability to regain sinus rhythm. Post-reperfusion myocardial tissue was analyzed for cardiomyocyte viability and structural preservation using histology and flow cytometry, and for RNA sequencing.

**Supplemental Figure 2.**
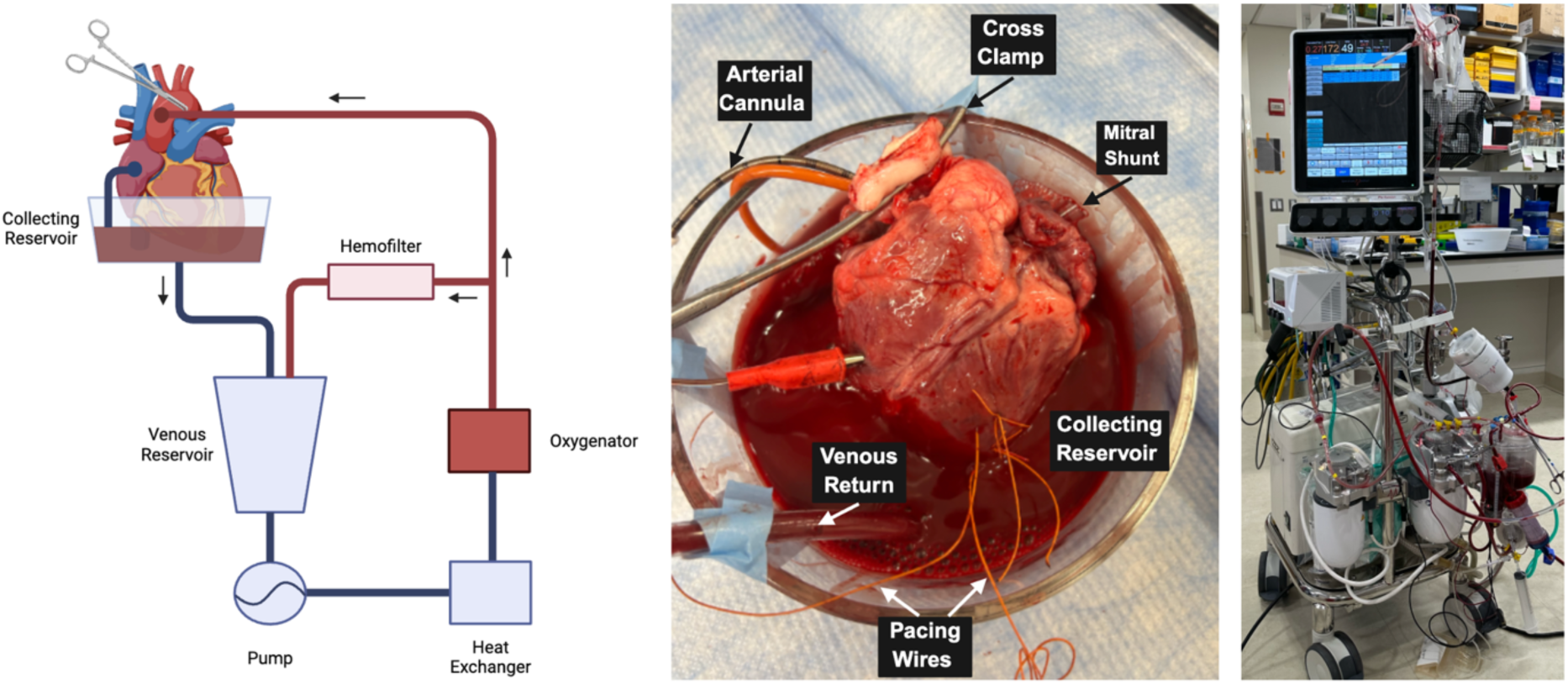
Schematic representation of normothermic machine perfusion circuit and photography of the NMP system. The NMP circuit consisted of a venous reservoir with Capiox FX05 oxygenator (Terumo Cardiovascular, Ann Arbor, MI), 3T Heater-Cooler system (LivaNova, London, UK), and Quantum 4-in. roller pump (Spectrum Medical, Gloucester, UK), and the circuit was primed with 500cc of whole blood from the donor pig. To load the heart onto the circuit, the ascending aorta was cannulated (8Fr Bio-Medicus NextGen Pediatric Arterial Cannula, Medtronic, Minneapolis, MN), and a silastic tube was maintained across the mitral valve to vent the left ventricle. The atria were widely opened, and venous return from the heart was allowed to passively drain into a collecting reservoir. The circuit was slowly initiated, the aorta de-aired, and an aortic cross-clamp was applied to begin 2 hours of reperfusion.

**Supplemental Figure 3.**
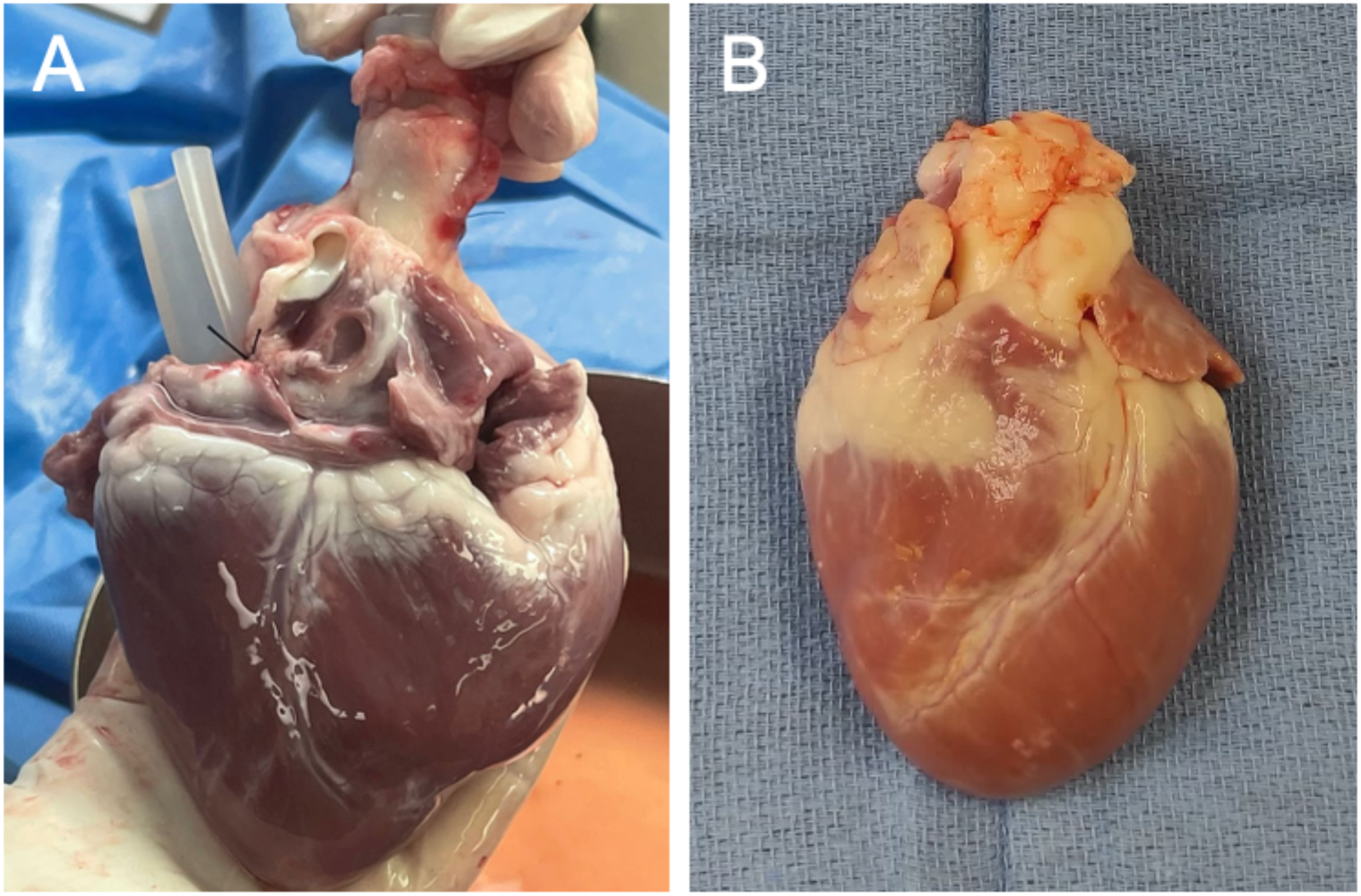
Gross morphology of porcine hearts following 48-hour *ex vivo* preservation. Representative images of porcine hearts preserved for 48 hours using either hypothermic oxygenated perfusion (HOPE) (Panel A) or static cold storage (SCS) (Panel B).

**Supplemental Figure 4.**
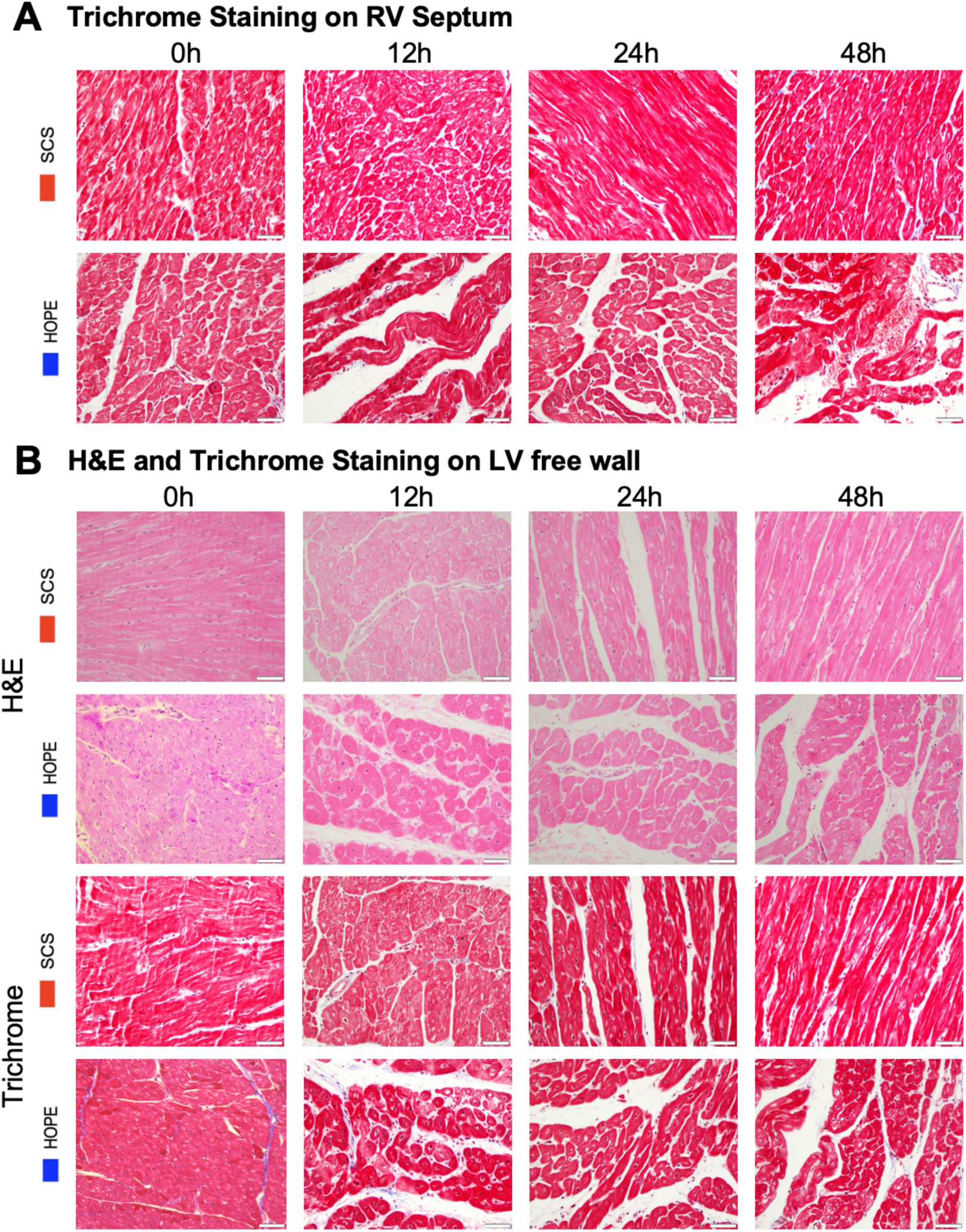
Histopathological assessment of cardiac tissue. **A)** Trichrome staining of RV septum of hearts preserved with HOPE or SCS at 0h, 12h, 24h, 48h. **B)** H&E and trichrome staining of LV free wall of hearts preserved with HOPE or SCS at 0h, 12h, 24h, 48h.

**Supplemental Figure 5.**
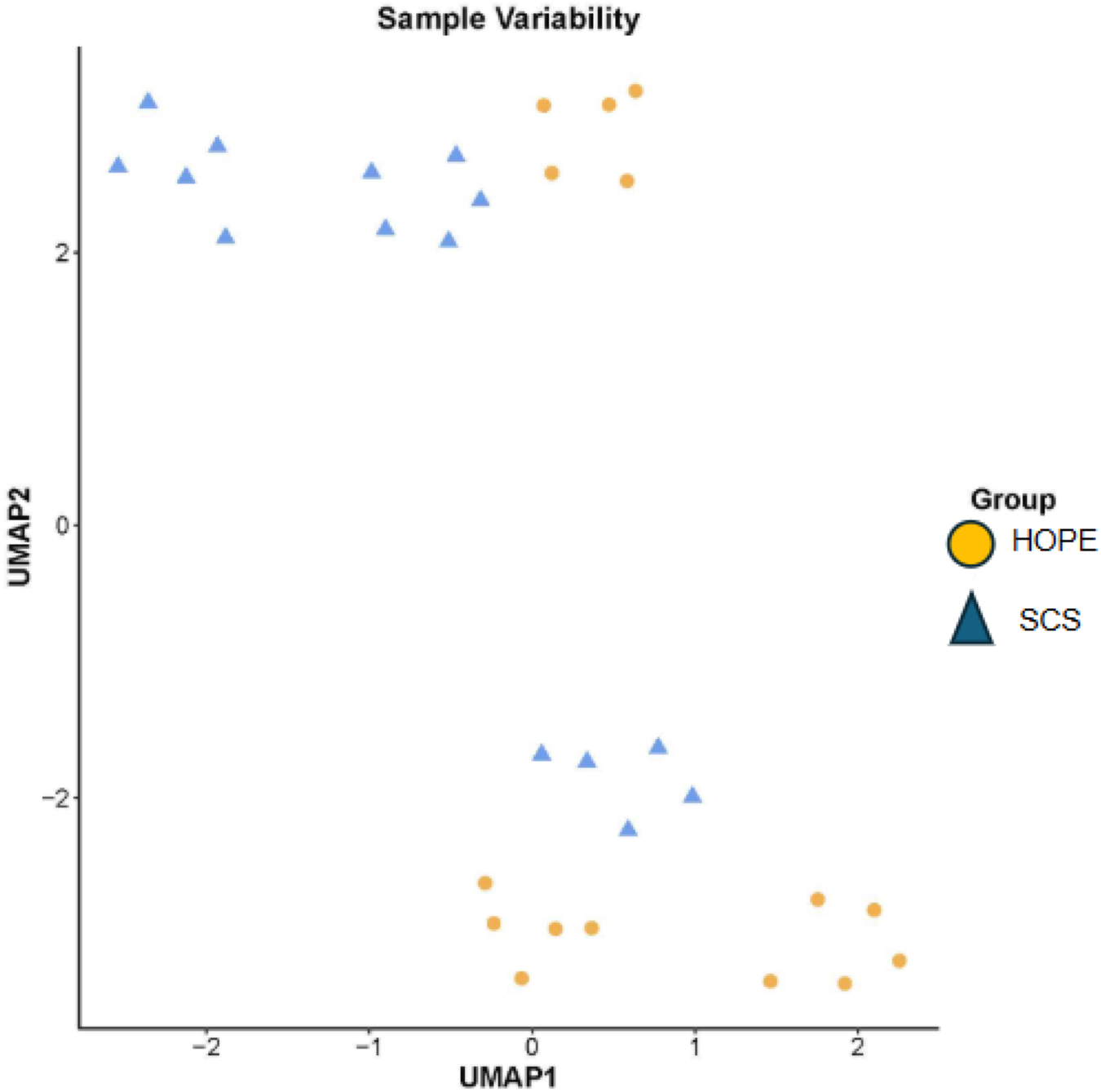
UMAP plot. Uniform Manifold Approximation and Projection (UMAP) provides a clearer representation of group variability within HOPE (yellow) and SCS (blue) samples.

**Supplemental Figure 6.**
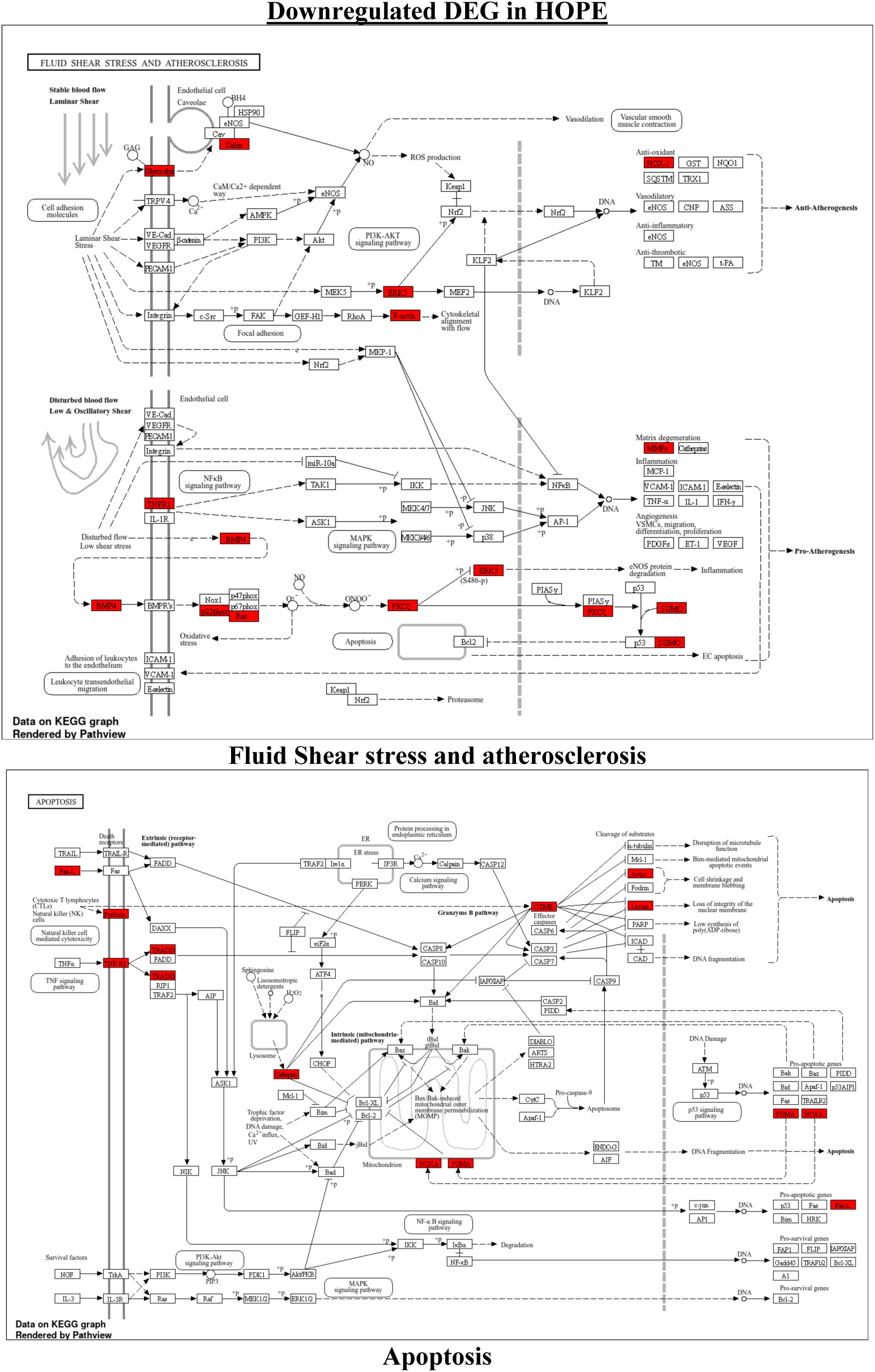
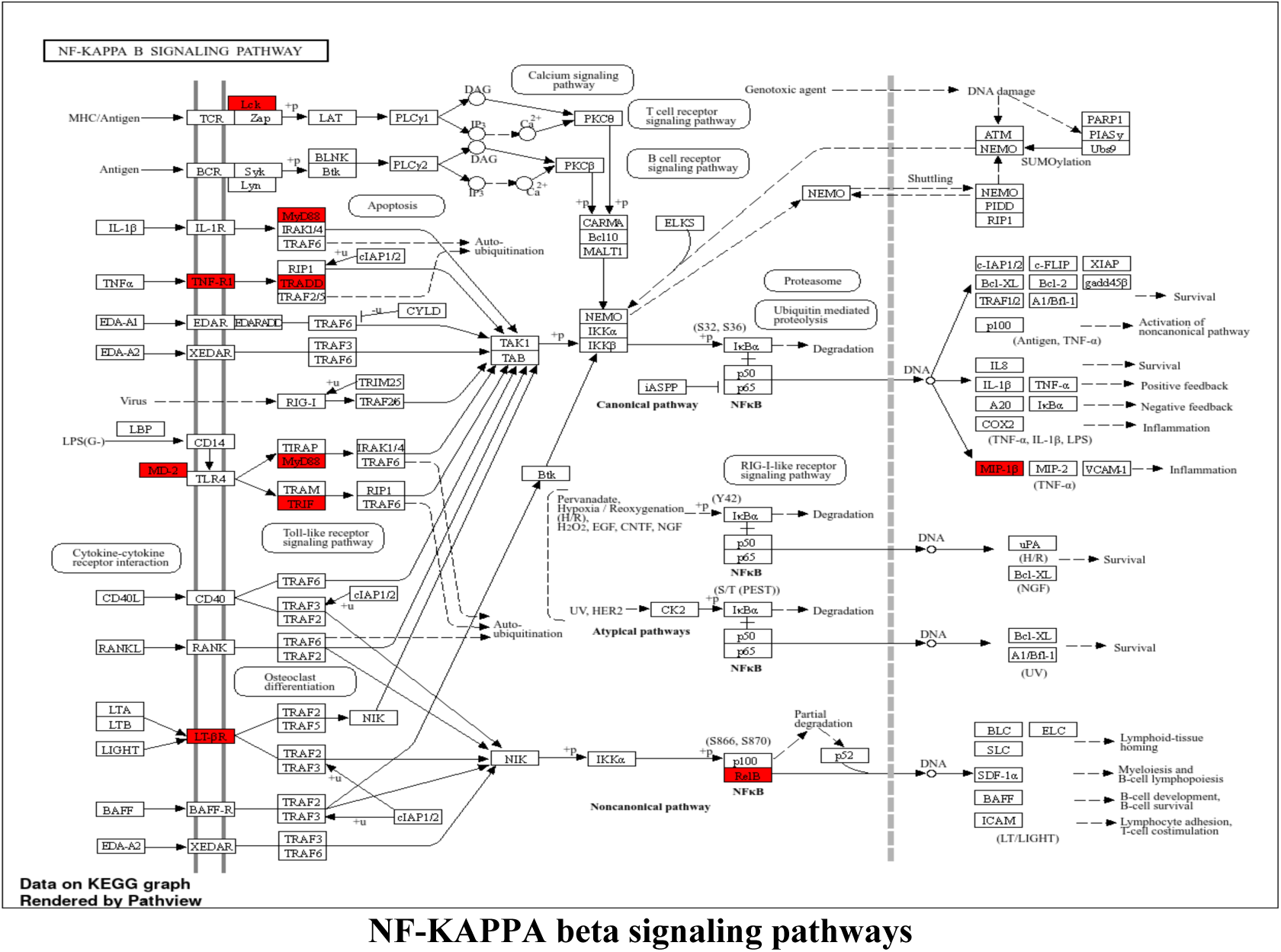
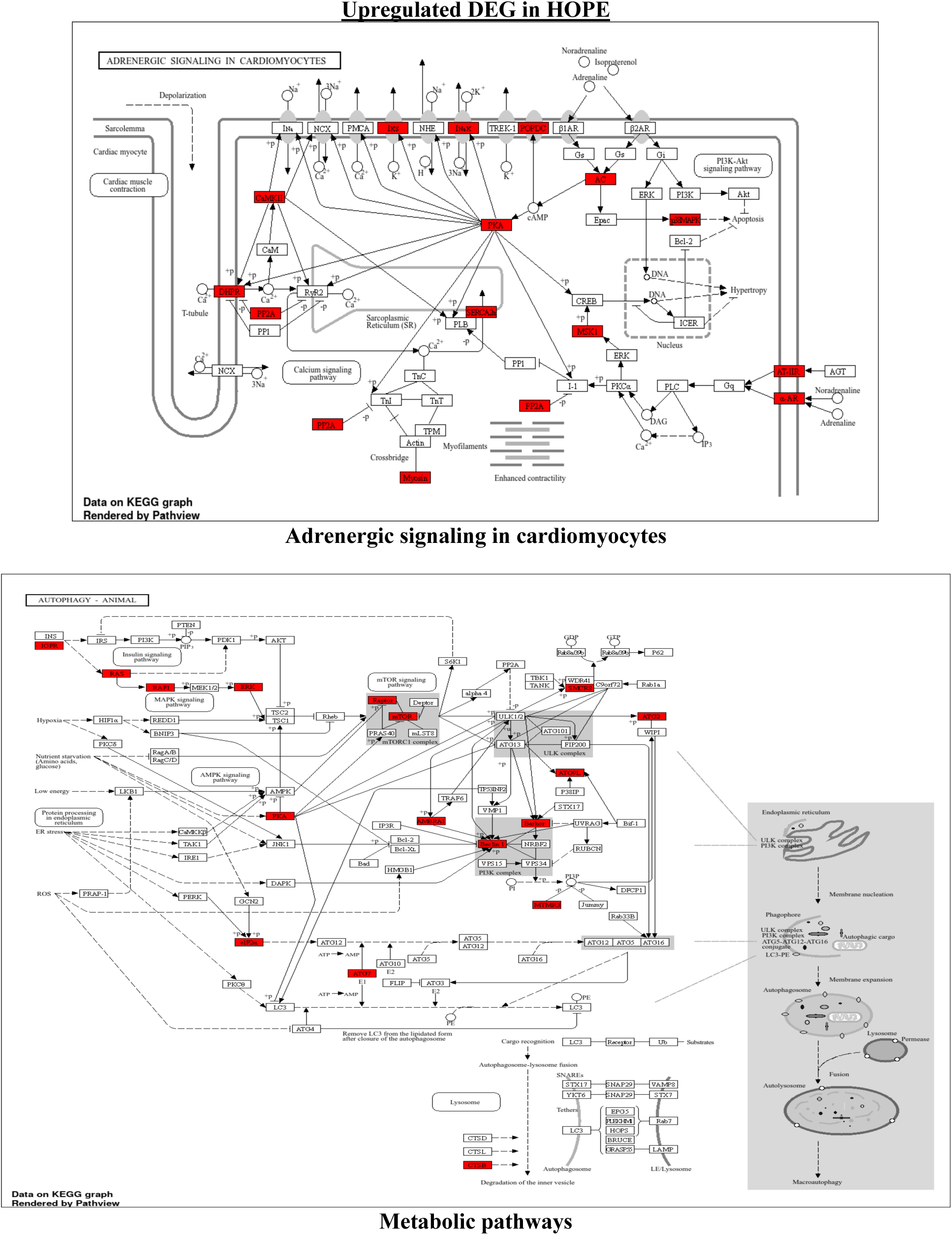
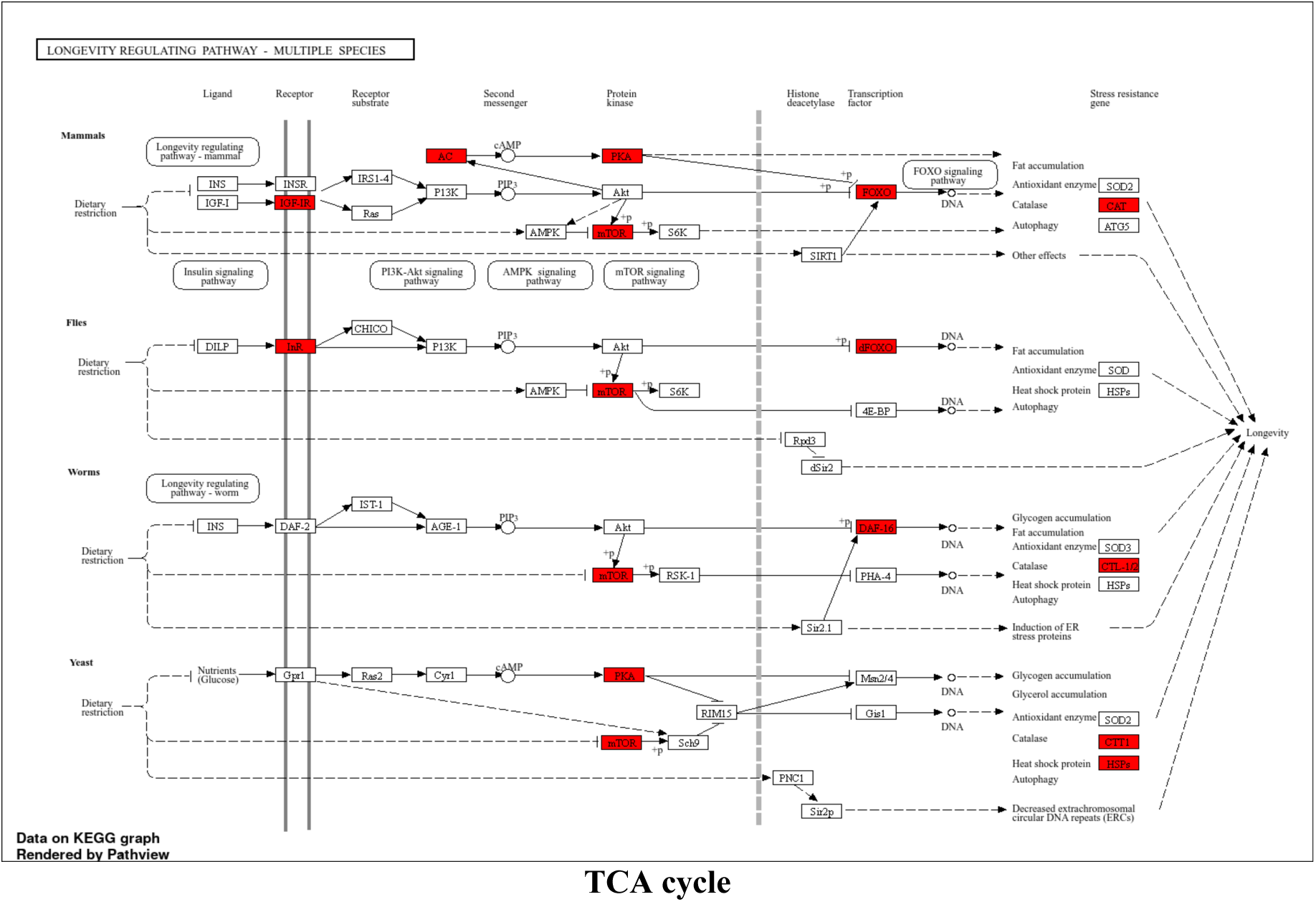
KEGG Pathway Mapping of Differentially Expressed Genes in HOPE-Preserved Hearts. Enriched KEGG pathways associated with differentially expressed genes (DEGs) in hearts preserved under hypothermic oxygenated perfusion (HOPE) at 24 and 48 hours. Pathways shown include key downregulated signals such as fluid shear stress and atherosclerosis, apoptosis, and the NF-κB signaling pathway, as well as significantly upregulated cardioprotective processes including adrenergic signaling in cardiomyocytes, core metabolic pathways, and the tricarboxylic acid (TCA) cycle. The diagram illustrates the distribution and functional roles of selected DEGs in each pathway, reflecting a global transcriptomic shift toward anti-apoptotic, anti-inflammatory, and metabolically active states in HOPE-preserved hearts. Color coding and arrows represent gene regulation status (upregulated in red, downregulated in blue) and directionality of signaling interactions.

**Supplemental Figure 7.**
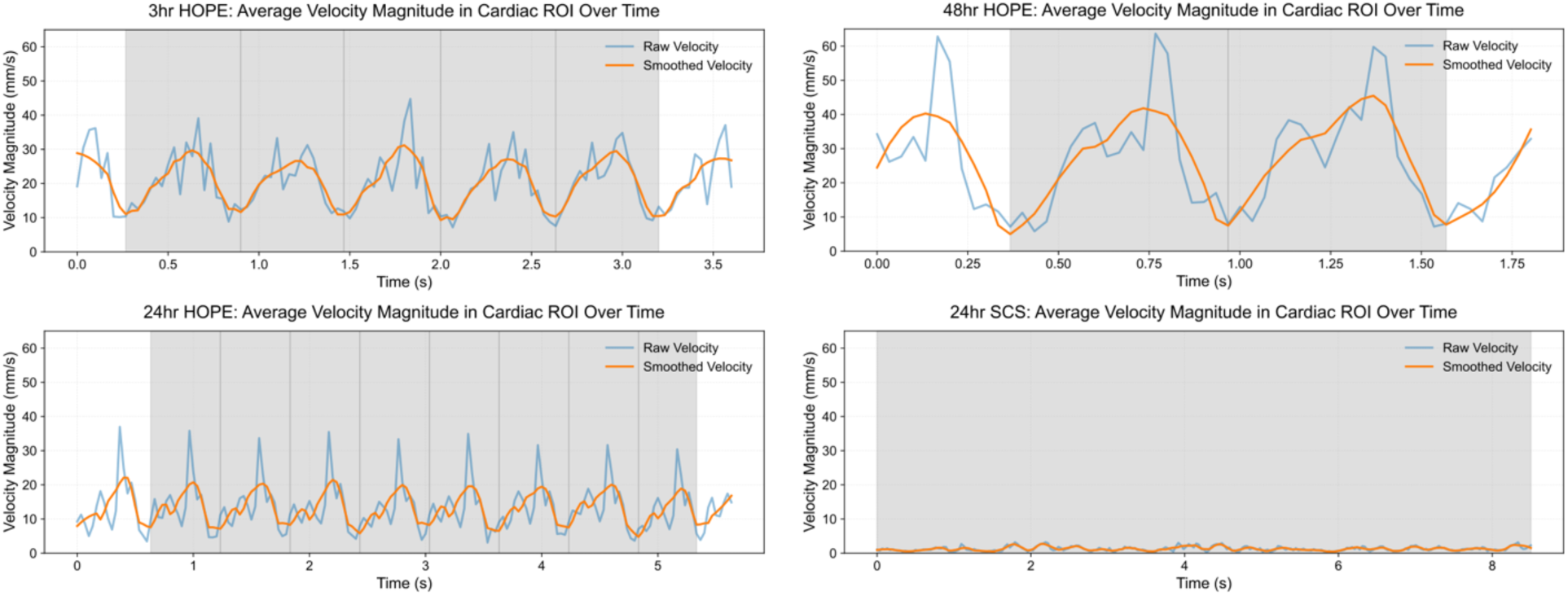
Mean Velocity Magnitude graphs during 2h of Normothermic Machine Perfusion following HOPE or SCS preservation. The velocity magnitudes (mm/s) derived from the frame-to-frame displacement of pixels across all contraction cycles were measured within a user-defined region of interest using the Farnebäck optical flow algorithm. The velocity signal was smoothed using a Savitzky-Golay filter (window length = 11, polynomial order = 2). HOPE-preserved hearts show high contraction velocities in each cycle, while SCS heart reports dramatically decreased velocity magnitude values, as it fails to regain sinus rhythm. The gray shading represents the data used for the analysis as it highlights all full cardiac cycles present. In the case of the SCS heart, where sinus rhythm was not regained, all timepoints were analyzed.

**Supplemental Figure 8.**
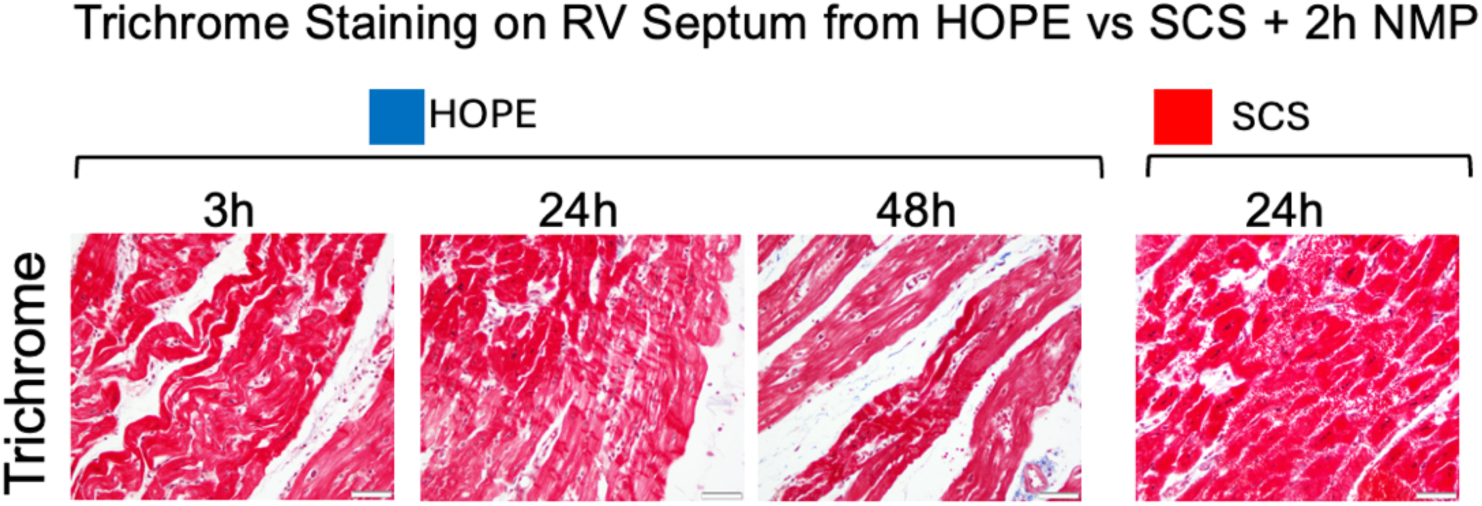
Trichrome Staining of RV Septum Following Normothermic Reperfusion in HOPE and SCS Hearts. Representative trichrome-stained histological sections from the RV septum of hearts preserved using hypothermic oxygenated perfusion (HOPE) for 3, 24, or 48 hours or static cold storage (SCS) for 24 hours, followed by 2-hour normothermic machine perfusion (NMP). HOPE-preserved hearts demonstrated preserved myocardial architecture with no evidence of interstitial fibrosis across all time points. In contrast, the SCS 24h heart showed disorganized tissue architecture and loss of structural integrity, consistent with severe reperfusion injury. These images reflect the structural outcomes observed in Arm 2 of the study assessing functional recovery post-reperfusion. Scale bars = 50 µm.

**Supplemental Figure 9.**
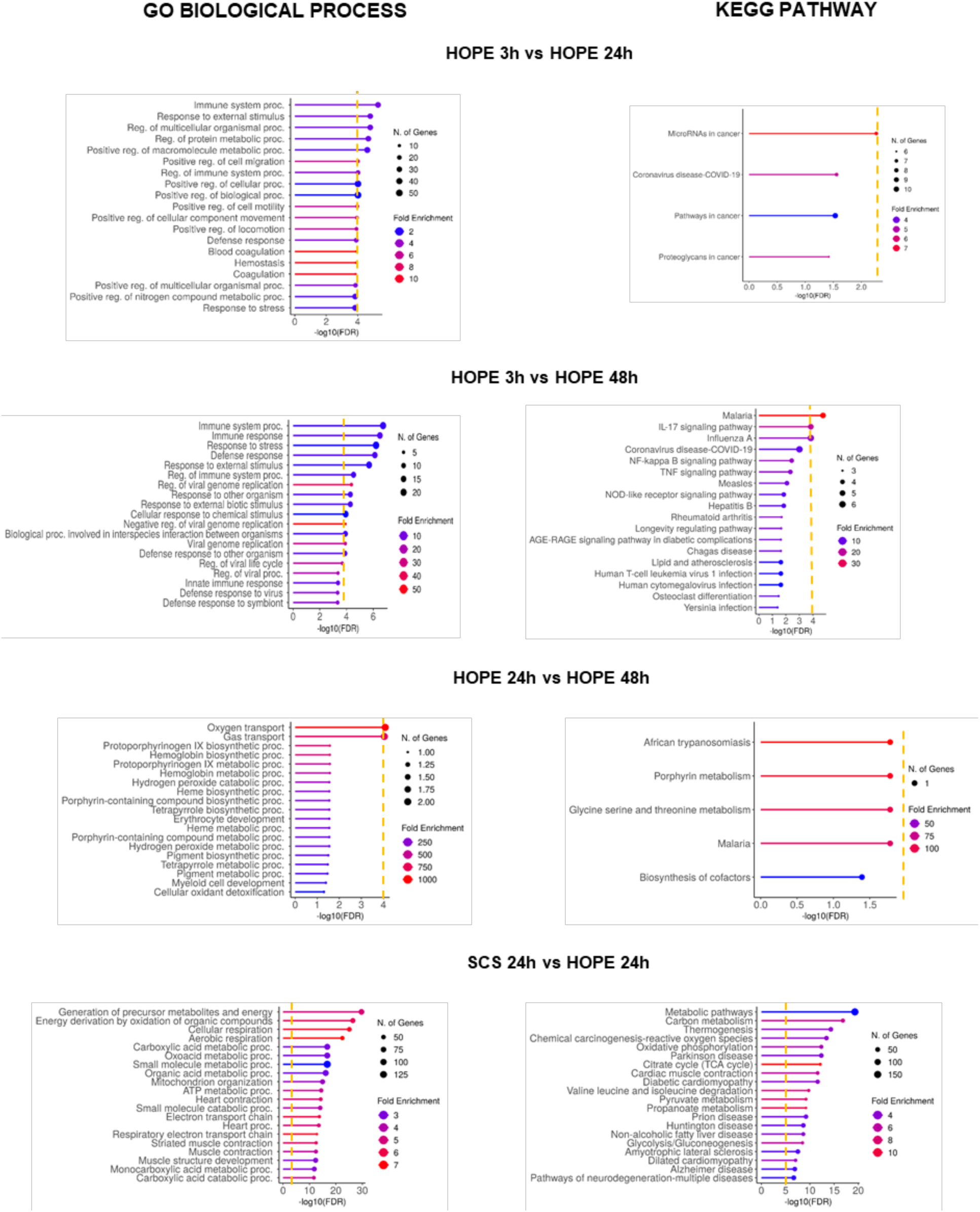
Gene ontology (GO) terms and KEGG pathway enrichment analysis of significantly upregulated genes in 3, 24, and 48h HOPE preservation groups. Dashed lines indicate minimum threshold of *FDR (False Discovery Rate) < 0.001* for less than 100 DEGs *and FDR < 10E-5* for more than 100 genes.

**Supplemental Figure 10.**
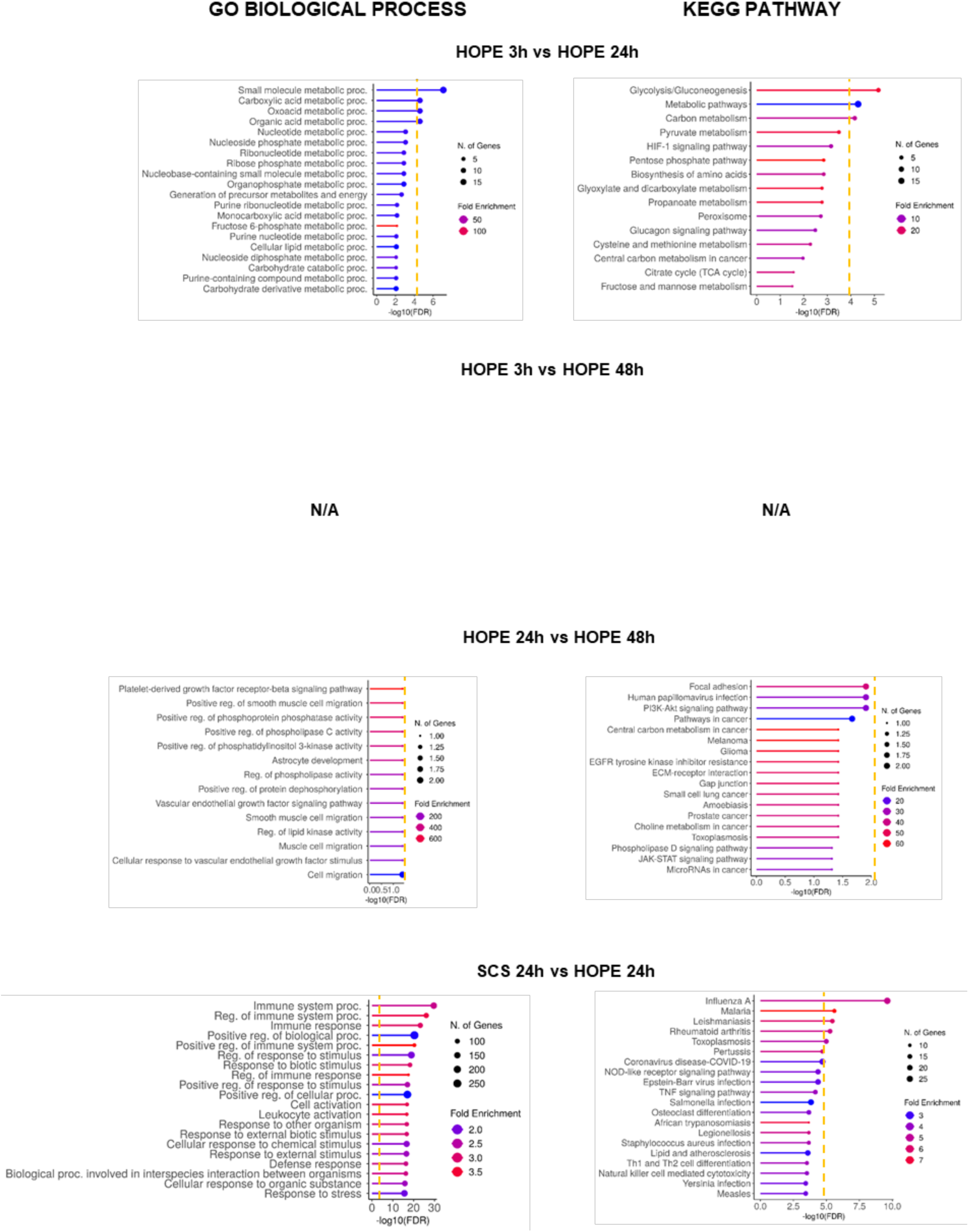
Gene ontology (GO) terms and KEGG pathway enrichment analysis of significantly downregulated genes in 3, 24, and 48h HOPE preservation groups. Dashed lines indicate minimum threshold of *FDR (False Discovery Rate) < 0.001* for less than 100 DEGs *and FDR < 10E-5* for more than 100 genes.

**Supplemental Table 1.**
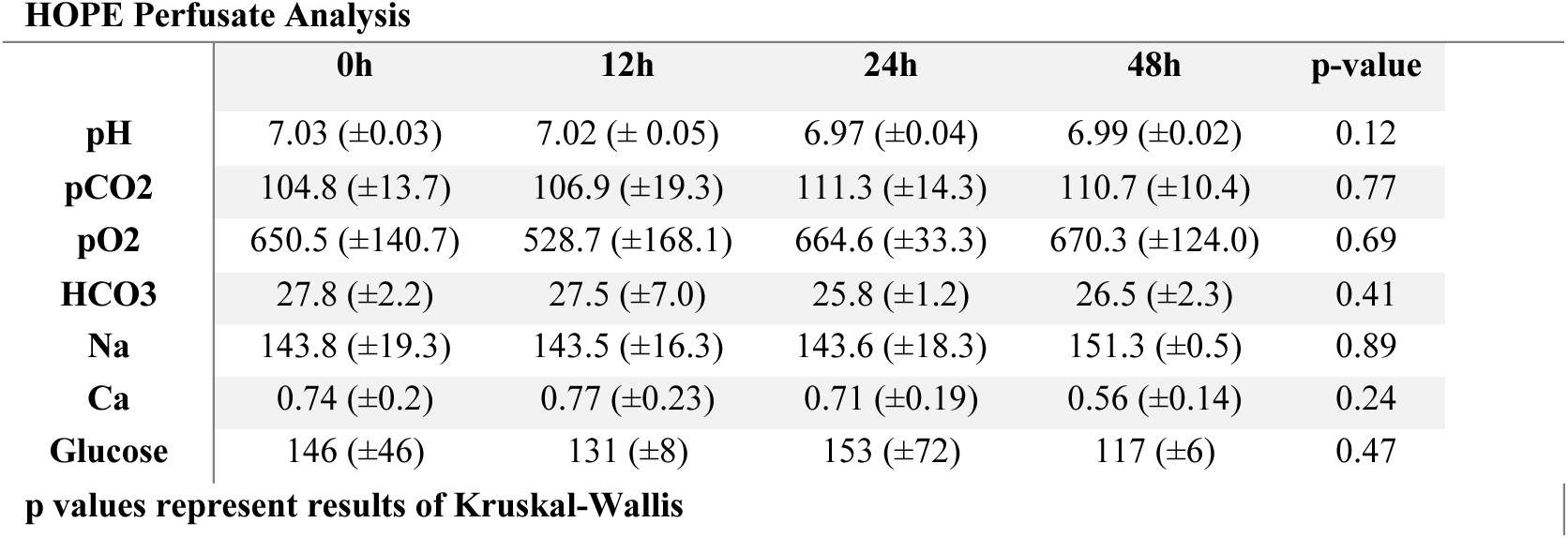
HOPE Perfusate Composition Over Time. Analysis of HOPE perfusate over time (0h, 12h, 24h, 48h). EPOC analysis of gas, electrolyte, and metabolite. Kruskal-Wallis test.

**Supplemental Table 2.** Pathway-Enriched Differential Gene Expression Profiles in HOPE vs. SCS Preservation. KEGG pathway maps highlighting gene-level transcriptional regulation in porcine hearts preserved with hypothermic oxygenated perfusion (HOPE) compared to static cold storage (SCS) at 24 and 48 hours. The diagram illustrates several key pathways altered in HOPE-preserved hearts, including oxidative stress, reactive oxygen species (ROS) signaling, apoptosis, mitochondrial function, and AGE/RAGE signaling. Genes significantly downregulated in HOPE (e.g., *S100A6, CTSW, SERPINE2, ND4, COX3*) suggest reduced oxidative injury, mitochondrial dysfunction, and apoptotic activity relative to SCS. In contrast, upregulated genes (e.g., *NLRX1, CDK5RAP1, LPL, EZR*) indicate activation of adaptive metabolic and stress response programs. Notably, EZR (ezrin) and S100A6 are involved in multiple pathways, reflecting integrated responses across signaling modules. Gene color coding corresponds to regulation direction (red = upregulated, blue = downregulated) and reflects transcriptomic comparisons at both 24h and 48h time points. Pathway overlay and gene-specific annotations were rendered using KEGG Mapper and manually curated to highlight the translational relevance of hypothermic oxygenated preservation.

| HOPE_VS_SCS_24H |  |  |  |
| --- | --- | --- | --- |
| PATHWAY | Gene Name | log2 FC | Adjusted p-value |
| OXIDATIVE STRESS-RELATED GENES | CDK5RAP1 | 1.38 | 5.89e-05 |
| OXIDATIVE STRESS-RELATED GENES | RAC2 | -1.38 | 5.69e-04 |
| OXIDATIVE STRESS-RELATED GENES | LGALS3 | -1.71 | 1.26e-03 |
| OXIDATIVE STRESS-RELATED GENES | STX2 | -0.93 | 1.43e-03 |
| ROS-RELATED GENES | CRY2 | 1.24 | 3.11e-04 |
| ROS-RELATED GENES | SLC2A4 | 1.18 | 3.60e-04 |
| ROS-RELATED GENES | CD44 | -0.86 | 1.38e-04 |
| ROS-RELATED GENES | S100A4 | -2.24 | 1.03e-03 |
| ROS-RELATED GENES | EZR | 1.25 | 9.99e-05 |
| ROS-RELATED GENES | NLRX1 | 1.12 | 1.30e-04 |
| ROS-RELATED GENES | H6PD | 1.03 | 9.08e-05 |
| APOPTOSIS-RELATED GENES | SERPINE2 | -1.77 | 1.38e-11 |
| APOPTOSIS-RELATED GENES | SUMO2 | -0.57 | 1.58e-03 |
| APOPTOSIS-RELATED GENES | EZR | 1.25 | 9.99e-05 |
| APOPTOSIS-RELATED GENES | PSMD2 | 0.64 | 2.13e-04 |
| APOPTOSIS-RELATED GENES | SLC2A4 | 1.18 | 3.60e-04 |
| APOPTOSIS-RELATED GENES | LPL | 1.37 | 2.51e-04 |
| APOPTOSIS-RELATED GENES | CD44 | -0.86 | 1.38e-04 |
| APOPTOSIS-RELATED GENES | S100A6 | -2.22 | 5.77e-04 |
| APOPTOSIS-RELATED GENES | CTSW | -1.59 | 1.00e-03 |
| APOPTOSIS-RELATED GENES | S100A4 | -2.24 | 1.03e-03 |
| APOPTOSIS-RELATED GENES | LGALS3 | -1.71 | 1.26e-03 |
| APOPTOSIS-RELATED GENES | GZMB | -2.88 | 1.45e-03 |
| MITOCHONDRIAL ACTIVITY | CTSW | -1.59 | 1.00e-03 |
| MITOCHONDRIAL ACTIVITY | S100A11 | -1.28 | 1.86e-05 |
| AGE/RAGE-RELATED GENES | EZR | 1.25 | 9.99e-05 |
| AGE/RAGE-RELATED GENES | S100A6 | -2.22 | 5.77e-04 |
| HOPE_VS_SCS_48H |  |  |  |
| PATHWAYS | Gene Name | Log2FC | Adjusted p-value |
| OXIDATIVE STRESS-RELATED GENES | CDK5RAP1 | 1.49 | 2.93E-06 |
| OXIDATIVE STRESS-RELATED GENES | SESN3 | -1.69 | 1.33E-10 |
| ROS-RELATED GENES | SERPINE2 | -1.35 | 7.91E-07 |
| ROS-RELATED GENES | NLRX1 | 1.66 | 8.47E-09 |
| ROS-RELATED GENES | GAA | 1.57 | 0.00012417 |
| ROS-RELATED GENES | COX3 | -2.43 | 1.68E-05 |
| ROS-RELATED GENES | SOX4 | -2.39 | 7.37E-05 |
| ROS-RELATED GENES | COX1 | -2.52 | 2.32E-07 |
| ROS-RELATED GENES | ATP6 | -2.44 | 8.60E-07 |
| ROS-RELATED GENES | S100A11 | -1.45 | 1.29E-07 |
| ROS-RELATED GENES | ND3 | -2.39 | 7.37E-05 |
| APOPTOSIS-RELATED GENES | EZR | 1.66 | 8.47E-09 |
| APOPTOSIS-RELATED GENES | PSMD2 | 0.80 | 2.46E-07 |
| APOPTOSIS-RELATED GENES | LPL | 1.54 | 1.03E-05 |
| APOPTOSIS-RELATED GENES | FOSL2 | 1.34 | 6.64E-05 |
| APOPTOSIS-RELATED GENES | BCL2L13 | 0.92 | 7.97E-05 |
| APOPTOSIS-RELATED GENES | SERPINE2 | -1.35 | 7.91E-07 |
| APOPTOSIS-RELATED GENES | S100A6 | -2.36 | 0.00012331 |
| MITOCHONDRIAL ACTIVITY | NLRX1 | 1.66 | 8.47E-09 |
| MITOCHONDRIAL ACTIVITY | CYTB | -2.38 | 8.90E-05 |
| MITOCHONDRIAL ACTIVITY | ND4L | -2.65 | 1.49E-10 |
| MITOCHONDRIAL ACTIVITY | ND4 | -2.43 | 1.44E-08 |
| AGE/RAGE-RELATED GENES | EZR | 1.66 | 8.47E-09 |
| AGE/RAGE-RELATED GENES | SCAP | 0.74 | 2.85E-07 |
| AGE/RAGE-RELATED GENES | S100A6 | -2.36 | 0.00012331 |
| AGE/RAGE-RELATED GENES | S100A11 | -1.45 | 1.29E-07 |

**Supplemental Table 3.** Differentially identified metabolites in HOPE vs SCS and their associated processes and pathways.

| PATHWAYS | METABOLITE | SCS_VS_HOPE_48H |  | SCS_48H_VS_0H |  | HOPE_48H_VS_0H |  |
| --- | --- | --- | --- | --- | --- | --- | --- |
|  |  | log2FC | p-value | log2FC | p-value | Log2FC | p-value |
| GLYCOLYSIS/<br>GLUCONEOGE<br>NESIS | AMP-Adenosine<br>5'-<br>monophosphate | 1.918880<br>088 | 0.00834<br>5636 | 0.4073642606 | 0.3023489<br>349 | -<br>1.980135936 | 0.01003546<br>397 |
| TCA CYCLE | Oleic Acid | 0.314726<br>694 | 0.20887<br>2692 | 0.4038391269 | 0.4002074<br>569 | 0.329818621 | 0.16203276<br>07 |
| TCA CYCLE | Flavin Adenine<br>Dinucleotide<br>(FAD) | -<br>0.516422<br>73 | 0.01828<br>4831 | -0.4826562684 | 0.0349805<br>9447 | -<br>0.047444877 | 0.73164463<br>78 |
| TCA CYCLE | Succinic Acid | -<br>0.538996<br>3411 | 0.29512<br>08787 | 0.04106051961 | 0.9348148<br>91 | 1.092727686 | 0.18487710<br>21 |
| ROS-RELATED<br>PATHWAYS | Arachidonic<br>Acid | -<br>1.181420<br>61 | 0.00813<br>5704 | -0.2900329408 | 0.2473225<br>42 | 0.790192921<br>1 | 0.00756457<br>9611 |
| ROS-RELATED<br>PATHWAYS | Hypoxanthine | 1.154090<br>039 | 0.17466<br>0686 | 0.5304349417 | 0.0392162<br>3288 | -<br>0.396920396<br>8 | 0.57416386<br>57 |
| ROS-RELATED<br>PATHWAYS | Glutamic Acid<br>(Glutamate) | -<br>0.102221<br>29 | 0.72676<br>0425 | -1.40232907 | 0.0107678<br>8871 | -<br>1.965476144 | 0.02080888<br>247 |
| ROS-RELATED<br>PATHWAYS | Glutathione<br>Reduced | 0.778098<br>949 | 0.48461<br>7103 | -0.7288715322 | 0.0242661<br>2865 | -<br>1.175231858 | 0.32602062<br>36 |
| ROS-RELATED<br>PATHWAYS | Oxidized<br>Glutathione | -<br>1.394430<br>752 | 0.07323<br>827997 | -2.055745046 | 0.0008245<br>78 | -<br>0.953149043<br>7 | 0.15092895<br>69 |
| ROS-RELATED<br>PATHWAYS | Eicosatrienoic<br>Acid | -<br>0.935349<br>49 | 0.05423<br>0572 | -0.7350729928 | 0.2668878<br>656 | 0.240319191<br>1 | 0.21628035<br>8 |

**Supplemental Video 1**: reanimation of heart during normothermic machine perfusion (NMP) after XVIVO (3h) preservation. <u>LINK</u>

**Supplemental Video 2**: reanimation of heart during normothermic machine perfusion (NMP) after XVIVO (24h) preservation. <u>LINK</u>

**Supplemental Video 3**: reanimation of heart during normothermic machine perfusion (NMP) after XVIVO (48h) preservation. <u>LINK</u>

**Supplemental Video 4**: reanimation of heart during normothermic machine perfusion (NMP) after SCS (24h) preservation. <u>LINK</u>

## Supplemental Material and Methods

### Contractility Quantification

#### Estimation of Pixel-to-Centimeter Conversion Factor

To quantify motion in physical units, a pixel-to-centimeter scaling factor was determined from a stabilized porcine heart video using OpenCV. A representative frame was extracted, and the user manually selected two points on the frame corresponding to a known 1 cm distance on the heart. The Euclidean distance in pixels between these two points was computed using the numpy.linalg.norm() function. This value was then used to calculate the conversion factor in pixels per millimeter. The scaling factor was subsequently applied to convert optical flow magnitudes from pixels to physical units.

#### ROI Selection and Optimization

The region of interest (ROI) used for motion quantification was defined as a rectangle. To ensure the ROI captured the largest possible area of the heart without including background artifacts, the center coordinates, dimensions, and angle of the rectangle were manually adjusted through iterative testing in the script. Several combinations were trialed, and the final parameters were selected based on visual alignment with the full beating region of the heart across frames.

#### Video Stabilization Using Dense Optical Flow

To correct for global motion artifacts caused by camera or table movement during heart perfusion, videos were stabilized using dense optical flow. Frame-to-frame displacements were estimated with the Farnebäck optical flow algorithm in OpenCV. The median flow vectors (dx, dy) were used to construct affine transformations that counteract global motion. These transformations were accumulated over time and applied to each frame to produce a stabilized output video. This preprocessing step focused the subsequent motion quantification on true tissue deformation rather than extrinsic movement.

#### Quantification of Velocity Magnitude from Optical Flow in a ROI

Stabilized perfusion videos of porcine hearts were analyzed using a custom Python pipeline to quantify myocardial motion and derive velocity magnitude metrics. Dense optical flow was computed between consecutive grayscale frames using the Farnebäck algorithm. Motion magnitude was averaged within a user-defined rectangular region of interest (ROI) and converted from pixels per frame to millimeters per second using a pre-calibrated spatial scale and video frame rate (30 fps). To suppress noise and preserve physiological waveform features, the velocity signal was smoothed using a Savitzky-Golay filter (window length = 11, polynomial order = 2). Systolic peaks and diastolic troughs were detected in the smoothed signal to extract the timepoints at which full cardiac cycles began and ended. If valid systole-diastole cycles were detected (i.e., meeting peak height and amplitude thresholds), mean velocity magnitudes across all full cardiac cycles were computed. If no valid cycles were found, the entire recording was treated as a single cycle and analyzed in full.

### Flow Cytometry

Fresh porcine cardiac tissue (0.2–0.3 g) was extracted using a 8mm surgical punch from the RV septum at designated time points. Biopsies were washed in PBS 1x (Corning) and then transferred to a petri dish containing warm enzymatic solution consisting of DMEM (Gibco) with 450 U/ml Collagenase I (Worthington, LS004196), 60 U/ml DNase I (Millipore, DN25) and 60 U/ml hyaluronidase (Worthington, LS002592). Tissues were minced into ∼2mm^3^ chunks then transferred into a 15 ml conical tube with the enzymatic solution and incubated in the rotating mixer (∼65/70 rpm) at 37 °C. After 1 h, the digestion was stopped by adding HBB buffer, consisting of 2% HI FBS (Gibco) and 0.2% BSA (Prometheus) in Hanks′ Balanced Salt solution (Sigma, H9269)^11^ filtered using a 100μm strainer. The suspended cells were centrifuged at 350g for 5 min at 4 °C, the supernatant was removed, and the pellet was resuspended with ACK lysing buffer (Gibco, A10492) to lyse red blood cells. After 5 min of incubation at room temperature, cells were washed with 9 ml of DMEM (Gibco), centrifuged at 350g for 5 min at 4 °C, and prepared for the flow cytometry. The cell suspension was blocked for 10 min on ice in blocking solution (PBS + 0.5% BSA). BSA was then washed out with PBS 1x washes. Cells were fixed in 2% PFA for 20 min on ice and washed twice in PBS1x. After fixation, cells were permeabilized with 0.1% Triton in PBS 1x 0.5% BSA for 20 min at 4°C^28^. Cells were stained with Alexa Fluor 647 mouse anti-cardiac troponin T (BD Bioscience – 565744) 1:50 for 1h on ice in blocking solution. DAPI was added (1:3000) and cells were analyzed with the NovoCyte Penteon Flow Cytometer Systems 5 Lasers (Agilent). Data were analyzed with NoveExpress 1.6.2 software. Controls: no-troponin staining. For compensation of fluorescence spectral overlap, UltraComp eBeads (eBioscience, Inc.) were used following the manufacturer’s protocols. The flow cytometry analysis was performed at the Columbia Stem Cell Initiative Flow Cytometry core facility at Columbia University Irving Medical Center.

### Whole Untargeted Metabolomics

To have a high-fidelity view of the whole metabolome, we performed a global untargeted metabolomic analysis of tissue samples collected during the experiments described above. Left ventricular biopsy samples were collected and flash frozen at various time points up to 48 hours of storage from both the HOPE and SCS preserved hearts (**Supplemental Fig.1A**). Untargeted Metabolomic profiling was conducted using ultrahigh performance liquid chromatography-high resolution accurate mass spectrometry (HRAMS) ^13^. Metabolites were extracted from tissue homogenates spiked with stable isotope labeled internal standards by protein precipitation using chilled 1:1 Methanol: acetonitrile^14^. Ten μl of the extract were analyzed using a high-resolution accurate-mass (HRAM) platform consisting of Vanquish™ Duo UHPLC system equipped with dual split sampler configuration coupled to a Exploris 240 HRAM mass spectrometer (Thermo Fisher Scientific, San Jose, CA, USA). Chromatographic separation was performed in triplicate using hydrophilic interaction liquid chromatography (HILIC) under positive ion mode and RP chromatography under negative ion mode, both at 60°C. HILIC separation will be done on a Waters XBridge BEH Amide XP column (2.1 mm, 50mm. 2.5 μm) and gradient elution with mobile phases 0.2% formic acid in water (A) and acetonitrile (B). Reverse-phase separation was performed on a C18 column (Higgins Targa C18 2.1 × 50 mm, 3 μm) with 1mM Ammonium acetate in water and acetonitrile as mobile solvents. The HRAMS was operated in a Full Scan (120K resolution) acquisition mode and a data dependent (DDA) acquisition mode with 60K in fullscan and 7500 ddMS2, both positive and negative polarity mode to acquire the spectral data.

Raw data files acquired through the Xcalibur software (version 4.1, ThermoFisher Scientific, MA, USA) were processed using the Compound Discoverer software (version 3.3.1, ThermoFisher Scientific, Waltham, MA, USA). The workflow used an adaptive curve model with 1 min maximum shift, 5 ppm mass tolerance, and 3 S/N (signal/noise) threshold for retention time alignment. Compound identification was achieved by using mzVault (internal ddMS2 database), mzCloud, and ChemSpider (using formula or exact mass and HMDB, KEGG, LipidMAPS as databases). Similarity searches for all compounds with ddMS2 data was done by using mzCloud and mzLogic algorithms applied to rank order ChemSpider results. For each identified metabolite, the null hypothesis was that there were no differences in area means across the groups that were tested using Student’s t-test and p-value, q-value and fold-change values for the comparisons were reported. Metabolites have been classified as whole feature list, all the detected signals from the mass spectrometer regardless of whether they have been identified as known metabolites, or annotated compounds list, corresponding to known metabolites that have been identified from the whole feature set using computational tools.

### RNA Sequencing

Isolation of total mRNA for RNA-seq was performed using the RNeasy mini kit (Qiagen, Valencia, CA). 1% β-mercaptoethanol was added to the lysis buffer and 20mg of previously flash-frozen frozen tissue were homogenized with a TissueRuptor (Qiagen). Isolated RNA purity was preliminarily assessed on a NanoDrop spectrophotometer (NanoDrop technologies). RNA concentration was measured on a Qubit 2.0 fluorometer (Invitrogen). RNA integrity assessment was performed on an Agilent Bioanalyzer 2100 using a Nano 6000 assay kit (Agilent Technologies) Columbia Genome Center core. Most of the samples presented an RNA integrity number (RIN) > 6. A total amount of > 600 ng RNA per sample was used as input material for the RNA sample preparations. Sequencing libraries were generated using Illumina TruSeq Stranded mRNA kit and modified the protocol by replacing the Illumina TruSeq PCR reaction with KAPA HiFi HotStart Ready Mix for the final PCR step to ensure compatibility with the 2×75bp paired-end sequencing on the Aviti Element. RNA sequencing was performed by the Columbia genomics core.

Bulk RNA-seq raw fastq files were first subjected to quality control using FastQCa (v0.12.1)^29^ to identify potential issues such as contamination and adapter sequences. After trimming low-quality sequences, FastQC was rerun on the cleaned reads to ensure that all major quality issues were resolved. The sequencing reads were then aligned to the Sus scrofa reference genome (Sscrofa11.1) using HISAT2 (v2.2.1)^30^. Gene expression levels were quantified with featureCounts (v2.0.6)^31^, and differential expression analysis was conducted using DESeq2^32^. The differentially expressed genes (DEGs) between different conditions were determined using the Wald test with a cut-off for absolute log2 fold change greater than 0.5 and adjusted p-values less than 0.05. Gene ontology (GO) analysis^33^ was conducted by using the ShinyGO application developed based on several R/Bioconductor functions. Finally, volcano plots and GO pathways were generated with the ggplot2 package.

## References

1. Jacob S, Garg P, Wadiwala I, et al. Strategies for Expanding Donors Pool in Heart Transplantation. Rev Cardiovasc Med. 2022;23(8):285. doi:10.31083/j.rcm2308285

2. Iyer A, Kumarasinghe G, Hicks M, et al. Primary graft failure after heart transplantation. J Transplant. 2011;2011:175768. doi:10.1155/2011/175768

3. Rega F, Lebreton G, Para M, et al. Hypothermic oxygenated perfusion of the donor heart in heart transplantation: the short-term outcome from a randomised, controlled, open-label, multicentre clinical trial. Lancet Lond Engl. 2024;404(10453):670–682. doi:10.1016/S0140-6736(24)01078-X

4. Kothari P. Ex-Vivo Preservation of Heart Allografts-An Overview of the Current State. J Cardiovasc Dev Dis. 2023;10(3):105. doi:10.3390/jcdd10030105

5. Nilsson J, Jernryd V, Qin G, et al. A nonrandomized open-label phase 2 trial of nonischemic heart preservation for human heart transplantation. Nat Commun. 2020;11(1):2976. doi:10.1038/s41467-020-16782-9

6. Steen S, Paskevicius A, Liao Q, Sjöberg T. Safe orthotopic transplantation of hearts harvested 24 hours after brain death and preserved for 24 hours. Scand Cardiovasc J SCJ. 2016;50(3):193–200. doi:10.3109/14017431.2016.1154598

7. See Hoe LE, Li Bassi G, Wildi K, et al. Donor heart ischemic time can be extended beyond 9 hours using hypothermic machine perfusion in sheep. J Heart Lung Transplant Off Publ Int Soc Heart Transplant. 2023;42(8):1015–1029. doi:10.1016/j.healun.2023.03.020

8. Brouckaert J, Dellgren G, Wallinder A, Rega F. Non-ischaemic preservation of the donor heart in heart transplantation: protocol design and rationale for a randomised, controlled, multicentre clinical trial across eight European countries. BMJ Open. 2023;13(12):e073729. doi:10.1136/bmjopen-2023-073729

9. Lebreton G, Leprince P. Successful heart transplant after 12 h preservation aboard a commercial flight. Lancet Lond Engl. 2024;403(10431):1019. doi:10.1016/S0140-6736(24)00258-7

10. Brouckaert J, Vandendriessche K, Degezelle K, et al. Successful clinical transplantation of hearts donated after circulatory death using direct procurement followed by hypothermic oxygenated perfusion: A report of the first 3 cases. J Heart Lung Transplant Off Publ Int Soc Heart Transplant. 2024;43(11):1907–1910. doi:10.1016/j.healun.2024.07.018

11. Koenig AL, Shchukina I, Amrute J, et al. Single-cell transcriptomics reveals cell-type-specific diversification in human heart failure. Nat Cardiovasc Res. 2022;1(3):263–280. doi:10.1038/s44161-022-00028-6

12. Ge X. DataMap: A Portable Application for Visualizing High-Dimensional Data. Published online April 11, 2025. doi:10.48550/arXiv.2504.08875

13. Liu Y, D’Agostino LA, Qu G, Jiang G, Martin JW. High-resolution mass spectrometry (HRMS) methods for nontarget discovery and characterization of poly-and per-fluoroalkyl substances (PFASs) in environmental and human samples. TrAC Trends Anal Chem. 2019;121:115420. doi:10.1016/j.trac.2019.02.021

14. Ramautar R, Berger R, van der Greef J, Hankemeier T. Human metabolomics: strategies to understand biology. Curr Opin Chem Biol. 2013;17(5):841–846. doi:10.1016/j.cbpa.2013.06.015

15. Lund LH, Khush KK, Cherikh WS, et al. The Registry of the International Society for Heart and Lung Transplantation: Thirty-fourth Adult Heart Transplantation Report-2017; Focus Theme: Allograft ischemic time. J Heart Lung Transplant Off Publ Int Soc Heart Transplant. 2017;36(10):1037–1046. doi:10.1016/j.healun.2017.07.019

16. Guo GR, Chen L, Rao M, Chen K, Song JP, Hu SS. A modified method for isolation of human cardiomyocytes to model cardiac diseases. J Transl Med. 2018;16(1):288. doi:10.1186/s12967-018-1649-6

17. Stokman G, Kors L, Bakker PJ, et al. NLRX1 dampens oxidative stress and apoptosis in tissue injury via control of mitochondrial activity. J Exp Med. 2017;214(8):2405–2420. doi:10.1084/jem.20161031

18. Mo L, Xu S, Jiang J, da Silva Menezes Júnior A, Huang D. Metabolic cardiomyopathy associated with a compound heterozygous variant in NAD(P)HX dehydratase: a case report and literature review. Transl Pediatr. 2024;13(12):2292–2304. doi:10.21037/tp-24-476

19. Schneider M, Kostin S, Strøm CC, et al. S100A4 is upregulated in injured myocardium and promotes growth and survival of cardiac myocytes. Cardiovasc Res. 2007;75(1):40–50. doi:10.1016/j.cardiores.2007.03.027

20. Tamaki Y, Iwanaga Y, Niizuma S, et al. Metastasis-associated protein, S100A4 mediates cardiac fibrosis potentially through the modulation of p53 in cardiac fibroblasts. J Mol Cell Cardiol. 2013;57:72–81. doi:10.1016/j.yjmcc.2013.01.007

21. Pulinilkunnil T, Rodrigues B. Cardiac lipoprotein lipase: metabolic basis for diabetic heart disease. Cardiovasc Res. 2006;69(2):329–340. doi:10.1016/j.cardiores.2005.09.017

22. Blondelle J, Lange S, Greenberg BH, Cowling RT. Cathepsins in heart disease-chewing on the heartache? Am J Physiol Heart Circ Physiol. 2015;308(9):H974–976. doi:10.1152/ajpheart.00125.2015

23. Kim EY, Zhang Y, Ye B, et al. Involvement of activated SUMO-2 conjugation in cardiomyopathy. Biochim Biophys Acta. 2015;1852(7):1388–1399. doi:10.1016/j.bbadis.2015.03.013

24. Kucera JA, Overbey DM, Turek JW. On-Table Reanimation of a Pediatric Heart from Donation after Circulatory Death. N Engl J Med. 2025;393(3):275–280. doi:10.1056/NEJMoa2503487

25. Wiig H, Swartz MA. Interstitial fluid and lymph formation and transport: physiological regulation and roles in inflammation and cancer. Physiol Rev. 2012;92(3):1005–1060. doi:10.1152/physrev.00037.2011

26. Vasques-Nóvoa F, Angélico-Gonçalves A, Alvarenga JMG, et al. Myocardial oedema: pathophysiological basis and implications for the failing heart. ESC Heart Fail. 2022;9(2):958–976. doi:10.1002/ehf2.13775

27. Michel SG, La Muraglia GM, Madariaga MLL, et al. Preservation of donor hearts using hypothermic oxygenated perfusion. Ann Transplant. 2014;19:409–416. doi:10.12659/AOT.890797

28. Waas M, Weerasekera R, Kropp EM, et al. Are These Cardiomyocytes? Protocol Development Reveals Impact of Sample Preparation on the Accuracy of Identifying Cardiomyocytes by Flow Cytometry. Stem Cell Rep. 2019;12(2):395–410. doi:10.1016/j.stemcr.2018.12.016

29. Andrews, Simon. FastQC: A Quality Control Tool for High Throughput Sequence Data [Online]. Published online 2010. http://www.bioinformatics.babraham.ac.uk/projects/fastqc/

30. Kim D, Paggi JM, Park C, Bennett C, Salzberg SL. Graph-based genome alignment and genotyping with HISAT2 and HISAT-genotype. Nat Biotechnol. 2019;37(8):907–915. doi:10.1038/s41587-019-0201-4

31. Liao Y, Smyth GK, Shi W. featureCounts: an efficient general purpose program for assigning sequence reads to genomic features. Bioinforma Oxf Engl. 2014;30(7):923–930. doi:10.1093/bioinformatics/btt656

32. Love MI, Huber W, Anders S. Moderated estimation of fold change and dispersion for RNA-seq data with DESeq2. Genome Biol. 2014;15(12):550. doi:10.1186/s13059-014-0550-8

33. The Gene Ontology Consortium. The Gene Ontology Resource: 20 years and still GOing strong. Nucleic Acids Res. 2019;47(D1):D330–D338. doi:10.1093/nar/gky1055

